# What innovations can address inequalities experienced by women and girls due to the COVID-19 pandemic across the different areas of life/domains: work, health, living standards, personal security, participation and education?

**DOI:** 10.1101/2022.05.04.22274659

**Authors:** Llinos Haf Spencer, Ned Hartfiel, Annie Hendry, Bethany Anthony, Abraham Makanjuola, Kalpa Pisavadia, Jacob Davies, Nathan Bray, Dyfrig Hughes, Clare Wilkinson, Deb Fitzsimmons, Rhiannon Tudor Edwards

## Abstract

**What is a Rapid Review?:** Our rapid reviews use a variation of the systematic review approach, abbreviating or omitting some components to generate the evidence to inform stakeholders promptly whilst maintaining attention to bias. They follow the methodological recommendations and minimum standards for conducting and reporting rapid reviews, including a structured protocol, systematic search, screening, data extraction, critical appraisal and evidence synthesis to answer a specific question and identify key research gaps. They take one to two months, depending on the breadth and complexity of the research topic/question(s), the extent of the evidence base and type of analysis required for synthesis.

**Background / Aim of Rapid Review:** The COVID-19 pandemic has led to differential economic, health and social impacts illuminating prevailing gender inequalities (WEN Wales, 2020). This rapid review investigated evidence for effectiveness of interventions to address gender inequalities across the domains of work, health, living standards, personal security, participation, and education.

**Key Findings:** *Extent of the evidence base:*

- 21 studies were identified: 7 reviews, 6 commentaries and 8 primary studies
- Limited evidence for the effectiveness of identified innovations in minority groups
- A lack of evaluation data for educational interventions
- A lack of evidence for cost-effectiveness of the identified interventions
- 14 additional articles were identified in the grey literature but not used to inform findings (apart from the Education domain, where there was a lack of peer-reviewed evidence).

*Recency of the evidence base:*

- All studies were published in 2020-2021

*Summary of findings:* Some evidence supported interventions/innovations related to ***work***:

- Permanent contracts, full-time hours, and national childcare programmes to increase income for women and thereby decrease the existing gender wage gap.
- More frequent use of online platforms in the presentation of professional work can reduce gender disparities due to time saved in travel away from home.

Some evidence supported interventions/innovations related to **health**:

- Leadership in digital health companies could benefit from women developing gender-friendly technology that meets the health needs of women.
- Create authentic partnerships with black women and female-led organisations to reduce maternal morbidity and mortality (Bray & McLemore, 2021).

Some evidence supported interventions/innovations related to **living standards** including:

- Multi-dimensional care provided to women and their children experiencing homelessness.

Limited evidence supported interventions/innovations related to **personal security** including:

- Specific training of social workers, psychologists and therapists to empower women to use coping strategies and utilise services to gain protection from abusive partners.
- Helplines, virtual safe spaces smart phone applications and online counselling to address issues of violence and abuse for women and girls.

Very limited evidence supported interventions/innovations related to **participation** including:

- Use of online platforms to reduce gender disparities in the presentation of academic/professional work.
- Ensuring equal representation, including women and marginalised persons, in pandemic response and recovery planning and decision-making.

Limited evidence from the grey literature described interventions/innovations related to **education** including:

- Teacher training curricula development to empower teachers to understand and challenge gender stereotypes in learning environments.
- Education for girls to enable participation in STEM.

*Policy Implications:* This evidence can be used to ***map against existing policies*** to identify which are ***supported by the evidence***, which are ***not in current policy and could be implemented*** and ***where further research/evaluation is needed*.** Further research is needed to ***evaluate the effectiveness of educational innovations,*** the ***effectiveness of the innovations in minority groups*** and the ***social value*** gained from interventions to address gender inequalities.

*Strength of Evidence:* One systematic review on mobile interventions targeting common mental disorders among pregnant and postpartum women was rated as high quality (Saad et al., 2021). The **overall confidence in the strength of evidence was rated as ‘low’ due to study designs**. Searches did not include COVID specific resources or pre-prints. There may be additional interventions/innovations that have been implemented to reduce inequalities experienced by women and girls due to the COVID-19 pandemic but have not been evaluated or published in the literature and are therefore not included here.

## 1. BACKGROUND

This Rapid Review is being conducted as part of the Wales COVID-19 Evidence Centre Work Programme. The above question was submitted by the Equality, Inclusion and Human Rights Branch of Welsh Government. The summary presented below will be considered by the Welsh Government Strengthening and Advancing Equality and Human Rights subgroup on Gender equality. It is intended to inform Welsh Government policy in reducing gender inequalities through interventions or innovations in the domains of work, health, living standards, personal security, and participation.

The COVID-19 outbreak brought unprecedented disruptions in patterns of work and childcare arrangements which have led to negative impacts on the mental health (O’Connor et al., 2021) and personal security of women (Ebert & Steinert, 2021). Homeworking and school closures (resulting in home schooling) have meant an increase in unpaid care work (Del Boca, Oggero, Profeta, & Rossi, 2021), which has highlighted gender inequality in the division of unpaid care work (WEN Wales, 2021). Women have been disproportionately impacted by the pandemic due to increased carer responsibilities and loss of income (Kyle, Isherwood, Bailey, & Davies, 2021; WEN Wales, 2020). Women in Wales were more likely to have lost their job due to a business closing down, with 18% of women experiencing job loss compared to 11% of men (Mohmed, 2021; WEN Wales, 2021).

COVID-19 is responsible for higher mortality in men. However, evidence suggests that COVID-19 has taken a greater toll on the mental wellbeing and physical health of women. During the first six weeks of lockdown in the UK (March to May 2020), UK research showed that women reported statistically significantly lower wellbeing than men (O’Connor et al., 2021). There is also evidence indicating that adolescent girls may have been among the most affected by emotional and psychological distress (Fong & Iarocci, 2020; McCluskey et al., 2021). The COVID-19 pandemic has also put women’s physical health and reproductive rights in jeopardy, as many countries such as Brazil, India, and Nepal reallocated their resources to the care of COVID-19 patients (United Nations, 2020). Such service closures are particularly concerning in countries where unsafe abortions are a leading cause of maternal death (Fisher & Ryan, 2021).

Globally, women earn less, save less, hold less secure jobs and are more likely to be employed in the informal and front-line care sectors that are prone to precarious employment conditions. In addition, women have inequitable access to social protection and head the majority of single-parent households. The aggregate result of these factors is a weakened ability to absorb adverse shocks such as the ones induced by the pandemic (United Nations, 2020). Evidence indicates some 47 million women will be pushed into poverty as a direct result of COVID-19. The threat of poverty has a disproportionate effect on women. In 2021, there were said to be 118 women in poverty for every 100 men. With this ratio threatening to widen further (Azcona et al., 2020), homelessness is a very real threat for many women due to the pandemic. To avoid this threat, some women may become trapped in dangerous living situations that put them at heightened risk of violence (Parker & Smith, 2021).

Among other concerning outcomes of the pandemic is the global rise in gender-based violence, including sexual assault and rape, experienced predominantly by women. Pooled data from around the world indicated that 45% of women reported that they or a woman they know experienced a form of violence against them since the start of the COVID-19 pandemic. 52% of unemployed women reported experience of violence against them, compared to 43% of employed women. One in two women with children experienced violence or knew a woman who had (Emandi, Encarnacion, Seck, & Tabaco, 2021). During the first week of lockdown in the UK, a 25% increase in the number of phone calls to the National Domestic Abuse Helpline was observed (UN Women, 2020). In their most recent review of the available literature, the Centre for Global Development (CGD) reported that 12 of the 15 papers discussing trends of pre-pandemic through intra-pandemic violence against women and children found evidence of increased violence. Loss of income and employment were identified as factors increasing the likelihood of violence (Bourgault, Peterman, & O’Donnell, 2021). Evidence has emerged on the make-up of this domestic violence, with abuse by current partners and family members having increased by 8% and 17% respectively within Greater London during the first lockdown (Ivandic, Kirchmaier, & Linton, 2020).

The negative effects are likely to be exacerbated for those from other marginalised groups, such as women from minority ethnic groups, women already living in poverty, and LGBT+ women, as these groups are at heightened risk of healthcare marginalisation, (Hafi & Uvais, 2020). All women and girls, including underserved populations, racial/ethnic or sexual minorities, immigrants and those with intersexual identities, will experience immediate and long-term consequences to their sexual and reproductive health as a result of the COVID-19 pandemic (Mukherjee et al., 2017). Women are underrepresented in national parliaments worldwide (making up only 25%). ∼For every one woman quoted in the media talking about the pandemic, there are three men (Freizer, Azcona, & Berevoescu, 2021) highlighting the underrepresentation and participation of women. Female participation is essential in emergency response groups but once again, women are underrepresented. This unequal participation in planning and decision-making roles put female-specific needs at risk of being overlooked (Freizer et al., 2021).

Overall, COVID-19 has exposed the impact of intersectionality, how characteristics such as class, sex, ethnicity, and disability combine to create interdependent and overlapping systems of discrimination, entrenching disadvantage. The COVID-19 pandemic has led to differential economic, health and social impacts illuminating prevailing gender inequalities (WEN Wales, 2020).

### 1.1 Purpose of this review

The purpose of this rapid review is to report on innovations or interventions that lessen gender inequalities in the domains of work, health, living standards, personal security, participation, and education.

The inclusion and exclusion criteria for included papers are presented below in Table 1. Although this rapid review was requested from stakeholders in the Welsh Government, studies from all OECD countries were included.

**Table 1:**
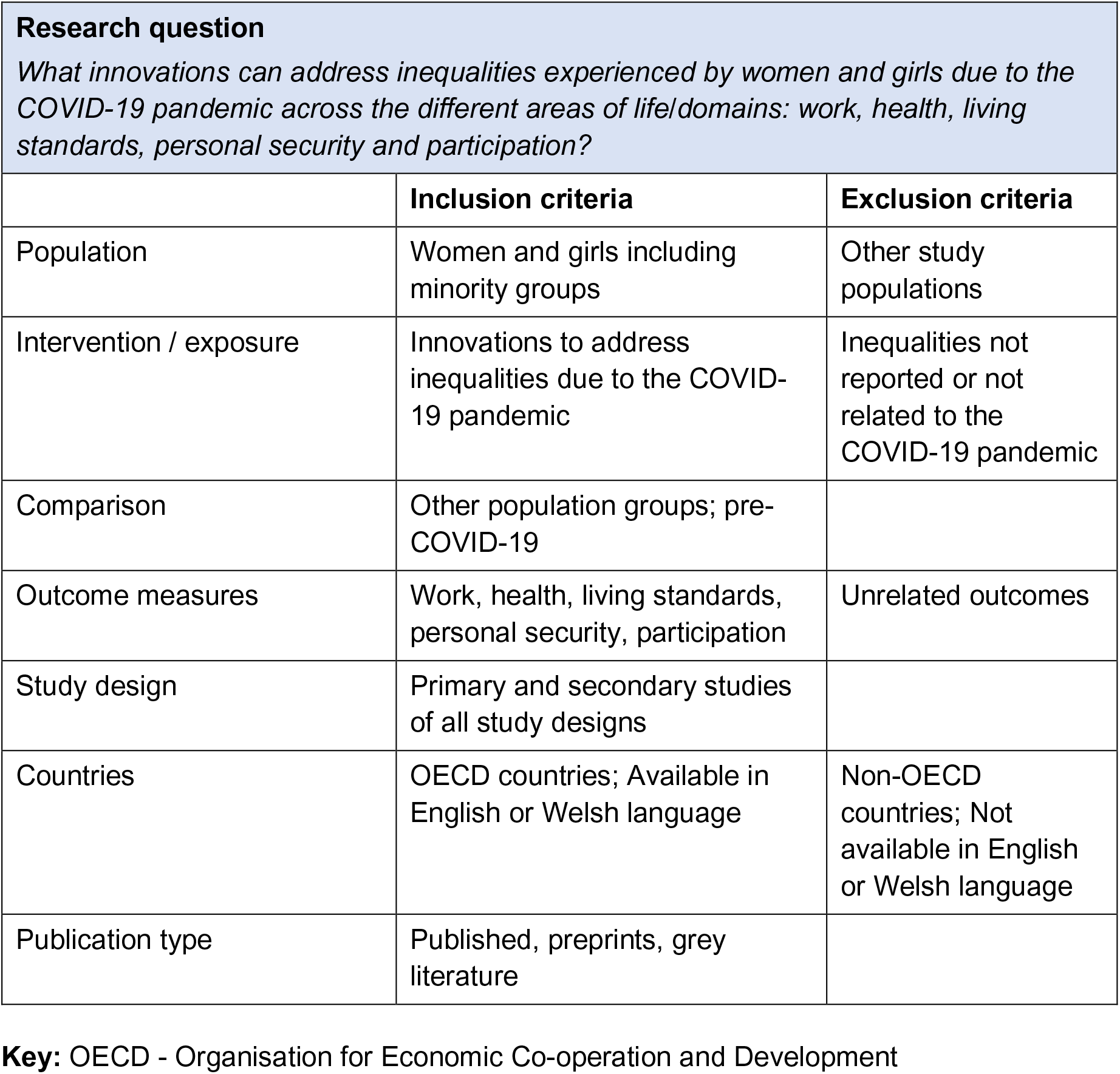
PICO and eligibility criteria.

## 2. RESULTS

The evidence collection methods are highlighted in section 6 of this report. However, in summary, 1291 studies were screened against title and abstract after identification of studies from six relevant databases. These were screened down to the 21 included papers (see Figure 1 for study selection flowchart). The included peer-reviewed papers (n = 21) on innovations/interventions to reduce gender inequality were synthesised to inform the results and mapped across six important ‘areas of life’ or ‘domains’ identified by Equality and Human Rights Commission (EHRC) of England, Wales and Scotland (see Table 2):

- work
- health
- living standards
- personal security
- participation
- education

**Table 2:** Number of studies per Equality and Human Rights Commission (EHRC) domain type.

| Evidence type | Work | Health | Living standards | Personal security | Participation | Education |
| --- | --- | --- | --- | --- | --- | --- |
| **Reviews** |  |  |  |  |  |  |
| **Systematic review** |  | Saad et al 2021 |  |  |  |  |
| **Rapid review** |  | Banati and Idele 2021 | Grammatikopoulou et al 2021 |  |  |  |
| **Scoping review** |  | Steinert et al 2021 | Andermann et al 2021 |  |  |  |
| **Scoping review** |  |  | Bray and McLemore 2021 |  |  |  |
| **Literature review** |  |  | Perri et al 2021 |  |  |  |
| **Review/reflection/qualitative discussion** | Moen et al 2021 |  |  |  |  |  |
| **Commentaries** |  |  |  |  |  |  |
| **Reflection/Commentary** | Banks et al 2021 |  |  | Hinton et al 2021 |  |  |
| **Viewpoint paper** |  | Figuro et al 2021 |  |  |  |  |
| **Concept paper** |  | Das et al 2021 |  |  |  |  |
| **Primary studies** |  |  |  |  |  |  |
| **Randomised controlled trial** |  |  |  |  |  |  |
| **Cross-sectional study** |  |  |  |  | Davic et al 2021 |  |
| **Cohort study** | Millan et al 2021 |  | Zhang et al 2021 |  |  |  |
| **Quantitative study** | Witteman et al 2020 | Ray et al 2021 |  | Chatzifotiou and Andreadou 2021 |  |  |
| **Qualitative study** | Kossek et al 2021 |  |  |  |  |  |
| **Case example/study** | Richey and Pointer 2021 |  | Goodsmith et al 2021 |  |  |  |
| **Total (n=21)** | **6** | **6** | **6** | **2** | **1** | **0** |

Fourteen additional articles were identified in the grey literature but are not included to inform the results section due to low quality/a lack of intervention evaluation data. Grey literature has been used to inform findings on educational interventions due to a lack of peer-reviewed research evidence in that area. Grey literature sources are tabulated in Appendix 2.

### 2.1 Six Equality and Human Rights Commission (EHRC) domains

**Work:** Six (n = 6) included papers were within the domain of work (Banks et al., 2021; Kossek, Dumas, Piszczek, & Allen, 2021; Millán, de la Torre, Rojas, & Jimber del Río, 2021; Moen et al., 2021; Richey & Pointer, 2021; Witteman, Haverfield, & Tannenbaum, 2021).

**Health:** Six (n = 6) included papers were within the domain of health (Banati & Idele, 2021; Das et al., 2021; Davic et al., 2021; Figueroa, Luo, Aguilera, & Lyles, 2021; Ray et al., 2021; Saad et al., 2021; Steinert, Alacevich, Steele, Hennegan, & Yakubovich, 2021)

**Living standards:** Six (n = 6) included papers were in the domain of living standards (Andermann et al., 2021; Bray & McLemore, 2021; Goodsmith, Ijadi-Maghsoodi, Melendez, & Dossett, 2021; Grammatikopoulou et al., 2021; Perri, Metheny, Matheson, Potvin, & O’Campo, 2021; Zhang, Limaye, & Means, 2021)

**Personal security:** Two (n = 2) included papers were within the domain of personal security (i.e., violence against women) (Chatzifotiou & Andreadou, 2021; Hinton et al., 2021).

**Participation:** One (n = 1) included paper was within the domain of participation (Davic et al., 2021)

**Education:** Although no (n = 0) included papers were within the domain of education, the grey literature (Appendix 2) indicated two relevant reports (Sands et al, 2021; Hammond et al, 2020).

### 2.2 Types of studies included

Although peer-reviewed papers (n = 21) and grey literature (n = 14) were investigated for inclusion, this rapid review focuses on the peer-reviewed papers summarised below according to six domains identified by Equality and Human Rights Commission (EHRC) of England, Wales and Scotland (apart from for the Education domain, where there was a lack of evidence in the peer-reviewed literature and so grey literature was drawn upon). The 21 peer-reviewed papers were a mixture of reviews, commentaries, and primary studies. Grey literature is included in Appendix 2.

**Reviews:** Seven (n = 7) included papers were reviews (Andermann et al., 2021; Banati & Idele, 2021; Bray & McLemore, 2021; Perri et al., 2021; Saad et al., 2021; Steinert et al., 2021) (see Table 3).

**Table 3.** Summary of review papers (n=7)

| Citation (Country) | Review details | Included studies | Quality assessment | Findings and observations/notes |
| --- | --- | --- | --- | --- |
| **Saad et al (2021)**<br>**Canada** | **Type of review:** systematic review<br>**Review period:** From inception of database to June 2020. Updated searches up in January 2021.<br>**Review purpose:** examine effectiveness and equity impact of mobile interventions targeting common mental disorders among pregnant and postpartum women<br>**Included study designs:** Quantitative, randomised and quasi- randomised controlled trials (RCTs; qRCTs), non-randomised controlled studies, controlled before and after studies and controlled interrupted time series.<br>**Quality rating:** High | **Number of included studies:** 18 studies included in quantitative analysis. 14 studies included in equity analysis.<br>**Key characteristics:** pregnant and postpartum women, women at any stage of pregnancy experience regardless of pregnancy outcomes. | **Primary findings:**<br>Prevention-based interventions showed potential to prevent depression and psychological distress.<br>For women in the antenatal stage, one study reported a statistically and clinically significant decrease in depression among the intervention group compared with usual care.<br>For women in the perinatal stage, two studies of peer support mobile applications showed statistically significant short-term improvements in depression severity.<br>For postpartum women, two studies reported that there was limited effectiveness on depression and anxiety symptoms. However, one study of a cognitive behavioural therapy mobile application showed a statistically significant and clinically important improvement.<br>Mobile interventions elicited significant improvements in mental health outcomes across East and South-East Asian ethnicities and across West-Asian ethnicities. | Mobile interventions refer to mobile phone features such as smartphone apps or text messages.<br>COVID-19 pandemic-related social isolation and uncertainty has increased and worsened levels of depression and anxiety in pregnant and postpartum women.<br>COVID-19 has increased barriers to accessing mental health care for this population.<br>Mobile interventions showed potential to reduce depression among antenatal and perinatal women. |
| **Banati & Idele (2021)** | **Type of review:** rapid review of reviews. | **Number of included studies:** | **Primary findings:** | Helplines, safe spaces, online |
| **Senegal and Italy** | **Review period:** 2000 to July 2020.<br>**Review purpose:** review literature on mental health outcomes and responses to HIV/ AIDS and identify lessons that could be used in response to COVID-19.<br>**Included study designs:** peer-reviewed, published systematic reviews.<br>**Quality rating:** Moderate | 63 systematic reviews drawing data from 2,498 studies.<br>**Key characteristics:** children and adolescents aged 0-19 years, all genders, and all HIV serostatus. | • The burden of care in the COVID-19 pandemic is falling largely on women and girls.<br>• Lockdown conditions led to a rise in domestic violence.<br>• Older adolescent girls, those with disabilities and victims of trafficking were at heightened risk of mental illness.<br>• Helplines, safe spaces, online counselling and virtual safe spaces may help address issues of violence and abuse for women and girls.<br>• Community-based mental health care for pregnant women may be helpful.<br>• Responses such as grants, food vouchers and community support groups may be helpful, although they would need to be adapted for COVID-19 and accommodate mask-wearing and social distancing. | counselling and virtual safe spaces may help address issues of violence and abuse for women and girls.<br>• Community-based mental health care for pregnant women may be helpful |
| **Steinert et al (2021)**<br>**Germany, UK, Australia, Canada.** | **Type of review:** rapid scoping review<br>**Review period:** 2005 to May 2020 and updated May 2021.<br>**Review purpose:** To review the evidence on gender-based interventions implemented in public health emergencies (PHEs).<br>**Included study designs:** all quantitative | **Number of included studies:** reporting on 16/20 relevant studies.<br>**Key characteristics:** women and girls. | **Primary findings relevant to OECD countries:**<br>• Reproductive health interventions were effective following the Zika virus outbreak in Puerto Rico, and reach was highest when messages were delivered via Facebook (Powell et al 2020).<br>• Four interventions consisting of telehealth or e-health approaches were implemented and scaled up in the context of the COVID-19 pandemic.<br>• One telehealth intervention showed greater uptake of virtual visits from HCPs among women (Darrat et al 2019). | Interventions were mostly implemented in low- and middle-income countries although 5 studies were implemented in the USA. Eight interventions were implemented in the context of natural disasters and the rest in the context of pandemics. These may be transferable to the COVID-19 pandemic. |
|  | **Quality rating:** Moderate |  | • A US study reported higher levels of satisfaction with virtual visits for pregnant women (Badri et al 2006).<br>• A telehealth intervention focused on prenatal care in Turkey reported significant reduction in distress and anxiety levels (Aksoy et al 2021). | • Three broad types of interventions were covered: economic empowerment, health promotion, and post disaster resettlement.<br>• Successful community support interventions need to be delivered with social distancing measures in place in the context of COVID-19.<br>• Virtual or telephone interventions may be more appropriate and effective where possible. |
| **Grammatikopoulou et al (2021)**<br>**Greece** | **Study Design:** Narrative review<br>**Review period:** April to May 2021<br>**Review purpose:** To present evidence of health challenges encountered by women experiencing homelessness.<br>**Included study designs:** Qualitative, quantitative and | **Number of included studies:** Unclear<br>**Key characteristics:** Water security<br>Stress<br>Food insecurity<br>Reproductive health<br>Aging<br>Mental health<br>COVID-19<br>Drug abuse | **Primary findings:**<br>• Transition to homelessness is often associated with poverty, unemployment, substance abuse, history of victimisation, stress, poor mental health and human immunodeficiency virus (HIV).<br>• Water insecurity can undermine bodily hygiene and dental health, posing a greater risk of dehydration and opportunistic infections.<br>• Exposure to extreme environmental conditions like heat waves and natural disasters increases morbidity, accelerates aging, and reduces life expectancy<br>• Food insecurity, obesity, and micronutrient deficiencies are apparent due to low diet quality and food waste. Poor hygiene, violence, and | • Care provided to women experiencing homelessness should be optimised in a multidimensional level<br>• Multi-dimensions include adequate safe water, sanitation access, psychological |
|  | mixed methods peer-reviewed studies. Grey literature including third sector and government reports and briefings, educational theses, conference proceedings.<br>**Quality rating:** Low | Food waste<br>Well-being<br>Sustainability | overcrowding increase the susceptibility of homeless women to communicable diseases, including sexually transmitted ones and COVID-19.<br>Established cardio-vascular disease and diabetes mellitus are often either undertreated or neglected, and their complications are more widespread in homeless women than in the general population.<br>Lack of medical screening and contraception non-use induce a variety of reproductive health issues. | support, disease management, acute health care, food of adequate quality, stress-coping strategies, opportunities for employment, support for minor dependents. |
| **Andermann et al (2021)**<br>**Canada** | **Study Design:** Scoping review<br>**Review purpose:** To identify evidence-based interventions and best practices to better support women experiencing or at risk of homelessness.<br>**Included study designs:** Quantitative and qualitative<br>**Quality rating:** Moderate | **Number of included studies:** 13 papers included, 4 systematic review and 9 randomised trials conducted in the USA (n=8), Netherlands (n=2), UK (n=1), Australia (n=1) and Canada (n=1).<br>**Key characteristics:** Equity<br>evidence-informed policy<br>gender<br>hidden homelessness<br>housing<br>intervention<br>research<br>scoping review<br>shelters; violence<br>women | **Primary findings:**<br>Interventions with the strongest evidence included:<br>post-shelter advocacy counselling for women experiencing homelessness due to intimate partner violence<br>case management and permanent housing subsidies (e.g., tenant-based rental assistance vouchers), which were shown to reduce homelessness, food insecurity, exposure to violence and psychosocial distress, as well as promote school stability and child well-being. | A population approach, involving greater investment in promoting gender equity, can improve outcomes for women and children before situations reach a crisis state.<br>Structural changes, are required including:<br>Formalising care work with pay scales and benefits<br>Transforming gender norms<br>Improving access to childcare<br>Ensuring pay equity<br>Creating opportunities for parental leave and work-life balance<br>Ensuring job protection for persons with<br>disabilities and other pro-equity policies and programs<br>• Helping to support families in creating stronger adult-adult and adult-child relationships<br>• Nurture social-emotional competencies for families through prenatal classes, nurse home visitation programs, day care facilities and schools to create greater family stability and to reduce intimate partner violence and adverse childhood experiences, which are often precursors to homelessness. |
| **Bray and Mclemore (2021)**<br>**USA** | **Study Design:** Scoping review<br>**Review period:** February 2021<br>**Review purpose:** To understand the experiences of black people and how scientific racism is manifest | **Number of included studies:** 67 papers<br>**Key characteristics:**<br>Black women<br>Black maternal health<br>White heteronormativity | **Primary findings:**<br>• Black women and pregnant capable people are not routinely consulted as experts on their own health<br>• Intervention studies are sparse and inadequate because they are focused on leveraging existing structures that are inequitable retrofits<br>• The bulk of scholarly contributions included reflect a lack of attention to black maternal health.<br>• Middle-class, white heteronormative gaze is more apparent in research questions and the determination of outcomes and exposures | • Gaps in data that do not account for realities of black life due to structural racism<br>• Social infrastructure inadequate to meet the health needs of black people. |
|  | in the conduct of clinical and public health research.<br>**Included study designs:**<br>Qualitative and quantitative.<br>**Quality rating:** Low |  | • Increase in black maternal death did not draw attention until the data began to indicate an increase in white maternal death.<br>• Much of the published research treats blackness as a universal characteristic, with little to no attention to the intersections of class, or income.<br>• Little or no attention to interventions specifically focused on low-income individuals and/or those using publicly funded insurance or services. | • Several action steps suggested:<br>○ Targeted Investments in black students, educators, healthcare providers, researchers<br>○ Targeted investments in social safety net and black communities<br>○ Put the voices, strategies, and interventions of black birthing people at the centre<br>○ Retrofit, reform, and reimagine clinical health services provision, education, research, and policy development<br>• Establish authentic partnership with black women and female led organisations as leaders of the work to reduce maternal morbidity and mortality<br>• Believe when interventions are<br>developed and implemented to improve black maternal health that the health of all populations will improve. |
| **Perri et al (2021)**<br>**USA** | **Study Design:** Literature review<br>**Review purpose:** To highlight how the exclusion of cisgender women and sexual and gender minorities (SGM) hampered recovery from the pandemic and further entrenched existing power structures that lead to the marginalisation of these groups.<br>**Included study designs:** Quantitative<br>**Quality rating:** Low | **Number of included studies:**<br>**Key characteristics:** COVID-19<br>Housing<br>Employment<br>Incarceration<br>Gender<br>Redistributive/transf<br>ormative policy | **Primary findings:**<br>• As well as physical distancing and lockdown policies, some health services that are essential for the health and wellbeing of sexual and gender minorities (SGM) people have been postponed, putting the health of these individuals at higher risk.<br>• Even before the pandemic, sexual and gender minorities (SGM) were more likely to lose employment (17%) compared with the general US population (13%) and less likely to have access to paid sick leave (29%) compared with the general population (76%)<br>• Women never recover pre-childbearing income levels even in countries with generous parental leave and job protection policies contributing to considerable wage losses over lifetimes.<br>• Applying an intersectionality lens reveals larger wage gaps by race/ethnicity or for being an SGM minority compared with white heterosexual men.<br>• In some countries, more than 80% of transgender women are thought to participate in sex work, making sex workers an especially important target population for gender redistributive/transformational employment and livelihood-related intervention in the context of COVID-19.<br>• Reports from Canadian violence against women shelters in 2018 demonstrated that 82% of individuals were turned away due to the shelters' limited capacity | • Lack of gender redistributive/transformational approaches in four sectors that rely on structural intervention for lasting change: health services, housing, employment, and incarceration<br>• Identifies how these sectors exacerbate various forms of oppression. |
|  |  |  | Since women (especially transgender women) represent a small proportion of the prison sector, they are largely invisible within a centralised, gender-blind and patriarchal system. |  |

**Abbreviations:** **SGM** - Sexual and Gender Minorities.**qRCT** – Quasi Randomised Controlled Trials

**Commentaries:** Six (n = 6) included papers were commentaries (Banks et al., 2021; Das et al., 2021; Figueroa et al., 2021; Goodsmith et al., 2021; Hinton et al., 2021; Richey & Pointer, 2021) (see Table 4).

**Table 4:** Summary of commentary papers (n=6)

| Citation (Country) | Review details | Included studies | Quality | Findings and observations/notes |
| --- | --- | --- | --- | --- |
| Das et al (2021)<br>USA | **Study Design:** Concept paper<br>**Type of intervention:** Virtual panel discussion<br>**Data collection methods:** Consensus development process via panel discussion<br>**Quality rating:** N/A | **Participants:** 70 members of the Academy for Women in Academic Emergency Medicine<br>**Setting:** Virtual<br>**Dates of data collection:** May 15<sup>th</sup> 2020 | **Primary findings:** 4 broad themes were highlighted and discussed by panellists.<br>Impact on clinical burden:<br>Women are more likely to hold teaching positions alongside clinical roles. Panellists reported an increase in teaching responsibilities, such as transferring curriculums to virtual platforms, with no reduction in clinical responsibilities leading to increased workload.<br>Work-life integration:<br>With additional pressures from COVID-19, many female physicians have found work-life integration even less tenable. Women hold greater burden of household responsibilities including, home schooling, cleaning, childcare, caring activities and food preparation.<br>Effects on academic productivity: | Whilst gender inequities in healthcare professionals were already in existence, the panel was asked to discuss whether they had been exacerbated by the pandemic.<br>The findings showed:<br>Women reported an increase in teaching responsibilities, such as transferring curriculums to virtual platforms, with no reduction in clinical responsibilities leading to increased workload.<br>Many female physicians found work-life integration less tenable. Women hold greater burden of household responsibilities including home schooling, cleaning, childcare, caring activities and food preparation.<br>Due to increased pressure from household and clinical work, female physicians found less time for research activity.<br>Women are less likely to hold executive leadership roles and due to the pandemic making in-person meetings was less possible. Academics and leaders have reverted to working with 'who they know' meaning women are even less visible. |
|  |  |  | Due to increased pressure from household and clinical work, time for research activity was often lost for female physicians.<br>4. Role of leadership and visibility:<br>Women are less likely to hold executive leadership roles, and due to the pandemic making in-person meetings less possible. Academics and leaders have reverted to working with 'who they know' meaning women are even less visible. |  |
| **Banks et al (2021)**<br>**Canada** | **Review period:** 2020 (No date specified)<br>**Review purpose:** identified challenges facing multidisciplinary faculty and trainees with dependents. Highlighted a number of possible strategies to address challenges in work-life (im)balance.<br>**Included study designs:** N/A - Policy recommendation document.<br>**Included outcome measures:** No outcome measures reported.<br>**Quality rating:** Low | **Number of included studies:** Not stated (policy overview)<br>**Key characteristics:** This paper highlights the issues faced by multidisciplinary faculty and trainees with dependents (i.e., children and elderly family members) in Canada. The authors outline potential strategies, with timelines and examples of implementation. | This policy recommendation document does not feature a study design. However, some innovations and strategies to reduce the effects of gender inequality for women include:<br>Ensure broad, equal representation (including women and marginalised persons) in pandemic response and recovery planning and decision-making<br>Canadian cardiovascular Society Equity and Diversity | This policy overview document offers several strategies for reducing the effects of gender inequality for women. However, no data was presented on the evaluation of these strategies. |
|  |  |  | Initiatives to improve mentoring and sponsorship of women and marginalised individuals.<br>• Canadian Institutes for Health Research implemented sex and gender training for researchers submitting grants.<br>• Canadian Institutes for Health Research targeted awards for sex- and gender-based analyses.<br>• Government funding of a national childcare program. |  |
| **Richey and Pointer (2021)**<br>**USA** | **Review period:** 2020 (No date specified)<br>**Review purpose:** To determine whether clinicians utilise an abbreviated cognitive behavioural therapy (CBT) model through an intersectional lens to augment the psychological well-being of women health-care workers (WHCW) in response to the pervasive stressors of the COVID-19 pandemic?<br>**Included study designs:** Case Study<br>**Included outcome measures:** PHQ-9 and GAD-7 scores | **Number of included studies:** One<br>**Key characteristics:**<br>• In a clinical case example, Trina, a 52-year-old, divorced mother of two, who worked full time as a nursing care manager, undertook a 4-session cognitive behavioural therapy model implemented through an intersectional lens. | • Cognitive behavioural therapy (CBT) is a therapeutic modality centered on teaching individuals to acknowledge and challenge distorted thinking patterns.<br>• The goal of psychotherapy through the lens of CBT is to increase the ability of the individual to create alternative, balanced thoughts to offset maladaptive thinking patterns. | • Investigated CBT as a therapeutic modality to use with women health-care workers in response to the pervasive stressors of the COVID-19 pandemic.<br>• CBT model requires empirical testing so it can be integrated in wider settings, specifically with women of colour to provide an 'anti-racist intersectional framework' |
|  | **Quality rating:** Moderate | The intervention was used to effectively decrease self-reported depressive and anxious symptomology via addressing aspects of gender inequality exacerbated through the COVID-19 pandemic.<br>These four sessions included CBT with a focus on gender-based equity and social justice and appeared to be beneficial for WHCWs during and potentially post the COVID-19 pandemic. |  |  |
| **Hinton et al (2021)**<br>**USA** | **Study Design:**<br>Reflection/commentary<br>**Purpose:** Aims to strengthen the evidence-base on multi-sectoral collaboration (MSC) research across intersecting services within health, education, social and financial protection, economic development, law enforcement to understand how they collaborate.<br>**Included study designs:** Qualitative and quantitative | **Number of included studies:**<br>**Key characteristics:**<br>Multi-sectoral collaboration<br>Sustainable development<br>Women<br>Children<br>Adolescents | **Primary findings:** This paper draws on a 12-country study series on MSC for health and sustainable development, in the context of the health and rights of women, children and adolescents.<br>Issues were analysed during the study period through 'real-time' discussions and structured reporting, as well as through literature reviews, surveys and retrospective feedback and analysis. | The authors of this reflection/commentary from the USA suggest several ways in which multiple sectors can work together to ensure appropriate care and support for female survivors of violence:<br>Reinforcing and extending emergency phones and 24-h hotlines and temporary shelters for survivors.<br>Adequately equipped first responders of violence against women to make prompt referrals to support services.<br>Security and justice sectors to promptly process complaints and protection orders.<br>Adjust security restrictions during the pandemic such as in Spain where women |
|  | **Quality rating:** Low |  | The authors identified four considerations that are unique to MSC research which will be of interest to other researchers, in the context of COVID-19 and beyond:<br>• Use theoretical frameworks to frame research questions as relevant to all sectors and to facilitate theoretical generalisability and evolution.<br>• Specifically incorporate sectoral analysis into MSC research methods<br>• Develop a core set of research questions, using mixed methods and contextual adaptations as needed, with agreement on criteria for research rigor<br>• Identify shared indicators of success and failure across sectors to assess MSCs. | who leave a situation of violence are exempt from lockdown.<br>• Partnering with communication and private sector providers to help expand technology-based solutions e.g., smart phone applications which can be used during lockdown restrictions to increase access to information on violence against women, service provision, and data collection. |
| **Goodsmith et al (2021)**<br>**USA** | **Study Design:** Case report<br>**Type of intervention:** Cross-agency collaboration in providing pregnant women and survivors of domestic violence with safe interim housing in the context of COVID-19.<br>**Data collection methods:** Case report<br>**Quality rating:** Low | **Participants:**<br>Unsheltered individuals who had survived domestic violence or who were pregnant.<br>**Setting:** 15,000 hotel rooms across Los Angeles County<br>**Dates of data collection:** During the first wave of the COVID- | **Primary Findings:**<br>• Local advocates successfully fought to add pregnant women to the list of high-risk populations eligible for private interim housing through the Project Roomkey Programme which pledged 15,000 hotel rooms to people with high-risk medical conditions during COVID-19 pandemic. | • This case report from the USA focused on the challenges of pregnant women and survivors of domestic violence who are experiencing homelessness.<br>• Rapid efforts and cross-agency collaboration in Los Angeles provided these groups with safe interim housing in the context of COVID-19. |
|  |  | 19 pandemic (March/April/May 2020). | With the support of a large private donation, the Mayor's Fund for Los Angeles launched Project Safe Haven to house DV survivors and their children in hotel rooms, with funding for up to 900 families. |  |
| **Figueroa et al (2021)**<br>**USA** | **Study Design:** Viewpoint paper<br>**Type of intervention:** Digital and telemedicine<br>**Data collection methods:** N/A<br>**Quality rating:** N/A | **Population:** Women in the USA<br>**Setting:** N/A<br>**Dates of data collection:** N/A | **Primary findings:**<br>The pandemic has exacerbated gender inequalities.<br>The use of digital health apps surged during the pandemic.<br>Digital health apps can boost gender equity through increased access to health information, yet digital health apps are rarely designed from a gender equity perspective.<br>Women are more likely to use health apps, but health apps are not always user friendly for women.<br>Digital health apps can place women at risk from partner violence and abuse if their phones are monitored.<br>Women with undocumented immigration status may also be wary of using apps which use location data. | Viewpoint paper illustrating gender issues in digital health from three levels: the users, the tools and the leadership<br>Offers recommendations for tackling gender bias through a feminist intersectional framework. |
|  |  |  | • Apps are often designed using gendered language, and apps for women are often stereotypically feminine and tend to focus on thinness and femininity.<br>• Apps often lack diversity, focusing on white, thin, young, middle-class women.<br>• Leadership in medicine and digital health companies is more likely to be male, leading to exclusion of women from development of technology that will target women. |  |

**Abbreviations:** Multi-sectoral collaboration (MSC).

**Primary studies:** Eight (n = 8) primary studies included five quantitative and three qualitative studies (see Table 5):

**Table 5:** Summary of primary studies (n=8)

| Citation (Country) | Study Details | Participants & setting | Key findings | Observations/notes |
| --- | --- | --- | --- | --- |
| **Davic et al 2021**<br>**USA** | **Study Design:** Cross sectional analysis<br>**Type of intervention:** N/A<br>**Data collection methods:** cross-sectional analysis of conference programs<br>**Quality rating:** Moderate | **Sample size:** 2,731<br>**Participants:** Speakers at national conferences for Emergency Medical Service (EMS) providers<br>**Setting:** National conferences in the USA<br>**Dates of data collection:** Various EMS conferences and trade shows in the United States (US) from 2016-2020. | **Primary findings:**<br>17 conference programs with 1,709 conference sessions identified for inclusion in the analysis.<br>Included sessions had a total of 2,731 listed speakers of whom 537 (20%) were female.<br>30 keynote addresses with 39 listed speakers of which six (15%) were female.<br>Changes to conference delivery due to COVID-19 may make presenting at conferences more accessible to women as no travel is required which may reduce non-attendance. | Cross-sectional analysis of conference presentations in the USA suggested that gender disparities in the presentation of academic/professional work could be reduced due to on-line platforms being used more often during the COVID-19 pandemic. |
| **Ray et al 2021**<br>**USA** | **Study Design:** Quantitative survey<br>**Type of intervention:** Online mental health and wellness intervention<br>**Data collection methods:** Online survey<br>**Quality rating:** Moderate | **Sample size:** 989<br>**Participants:** University students<br>**Setting:** A large University in the USA<br>**Dates of data collection:** June-September 2020 | **Primary findings:**<br>Gender was significant with men having more normal levels of anxiety and women having more moderate and severe anxiety.<br>Women reported greater levels of COVID-19 related worry than men.<br>Women experienced a greater number of daily life disruptions.<br>The greatest differences between men and women were difficulty in sleeping, loss of income and reduction in work hours. | Quantitative survey explored how students responded to COVID-19 and their use of an online wellness intervention.<br>Survey also assessed which student groups were in need of access to mental health support. |
| **Millan et al 2021**<br>**Spain** | **Study Design:** Quantitative survey investigating Spanish labour market with a special emphasis on women to determine how this market has evolved and the situation of the female | **Sample size:** Sample size not reported.<br>**Participants:** Men and women in the Spanish Labour Market.<br>**Setting:** The Spanish Labour Market | **Primary Findings:**<br>More stable jobs are advocated (to increase the percentage of permanent contracts and complete working days) that will favour an increase in income for | Quantitative survey investigating labour market trends in Spain over a 12-year period.<br>Findings advocated more stable jobs for women to |
|  | Spanish worker.<br>**Type of intervention:** The Spanish labour market was analysed through the labour force survey from a gender perspective.<br>**Data collection methods:** Labour Force Survey and analysis/ ARIMA analysis<br>**Quality rating:** Moderate | **Dates of data collection:** The data was collected from the first quarter of 2008 to the third quarter of 2020. | women, decreasing the existing wage gap.<br>Recommended reducing wage inequalities by setting a minimum wage which is the same for men and women.<br>Offer the same promotion to men and women to reduce differences between genders in high-rank positions.<br>Increasing the number of women in managerial occupations via quotas is not recommended. When quotas occur, the priority is to get women to fulfil the quota, which may mean hiring women who are not qualified.<br>Future research should include investigating the women's labour market according to types of enterprises (public or private) to compare levels of gender inequality between the private and public sectors. | increase the percentage of permanent contracts and complete working days.<br>Permanent contracts and full-time hours would increase the incomes of women, decreasing the existing wage gap.<br>The wage gap problem is not exclusive to Spain. |
| **Witteman et al 2020**<br>**Canada** | **Study Design:** Quantitative study<br>CIHR created a guidance document: 'Why sex and gender need to be considered in COVID-19 research' which and required reviewers to evaluate the integration of sex, gender, and other identity factors (e.g., age, race, ethnicity, culture, religion, geography, education, disability, income, and sexual orientation) at all stages of the research process. | **Sample size:** The first grant competition funded 100 of 227 applications (overall success rate 44%)<br>The second grant competition garnered greater application pressure, funding 139 of 1,488 applications (overall success rate 9%).<br>**Participants:** Those that self-identify as male, female, indigenous, disabled, gender fluid, non-binary, and 2S.<br>**Setting:** All PIs were based in Canada and made their submissions online.<br>**Dates of data collection:** | **Primary Findings:**<br>Following target gender policy changes, the funder received more applications from female scientists, awarded a greater proportion of grants to female compared to male scientists.<br>Funder received and funded more grant applications that considered sex and gender in the content of COVID-19 research.<br>Four observations were made:<br>Quality of submissions prioritised over speed.<br>Better to be more proactive than reactive with policy implementation.<br>The changes implemented by CIHR appeared to primarily solve problems | Requiring grant applications to consider sex and gender in COVID-19 research encouraged female researchers to submit more applications and be awarded more research grants. |
|  | **Type of intervention:**<br>Grant applications and success rates were compared from submissions by PIs with different identity characteristics.<br>Grant applications from PIs were examined for whether the applications accounted for sex and gender.<br>**Data collection methods:**<br>Data from grants applications was routinely collected when applicants create accounts in the online CIHR system.<br>Self-identify as female or male or did not provide an entry in that field.<br>**Quality rating:** Moderate | Grant competition 1 occurred in February 2020. Grant competition 2 occurred from April to May 2020. | related to applicants' sex and gender. Barriers related to other identity dimensions (race and ethnicity, Indigenous identity, disability) require further analysis and consultation to identify solutions.<br>4. Educational support and explicit evaluation criteria related to sex, gender, and other identity characteristics may help ensure that applicants and peer reviewers attend to these factors within the proposed research, thereby expanding definitions of research excellence.<br>People who self-identified as being part of a visible minority community were 30% of applicants in both COVID competitions, and 28% and 26% of funded investigators in the first and second competitions, respectively.<br>The numbers of applicants who identified as indigenous, disabled, gender fluid, non-binary, or Two-Spirit were too low (counts under five in submitted applications, funded applications, or both) to report in a disaggregated way.<br>Biological sex factored into 55% and 94% of funded applications in the first and second competitions, respectively.<br>In the second competition, applications indicating that sex was considered in the proposed work were more likely to be funded (odds ratio [OR] 3.13, 95% CI 1.57 to 6.23). This was not the case in the first competition (OR 1.31, 95% CI 0.91 to 1.88). |  |
| **Zhang et al (2021)**<br>**USA** | **Study Design:** Cohort study<br>**Type of intervention:** Evaluation of a collaborative care model between a tertiary care hospital, Brigham Women's Hospital (BWH), and a community organisation, Health Care without Walls, that bridges gaps in care for housing insecure, pregnant women.<br>Reimagine the care delivery system to meet the needs of housing insecure, pregnant women during the COVID-19 pandemic.<br>**Data collection methods:** Bridges to Moms (BTM) was evaluated using maternal and new-born chart data and tracking non-profit spending.<br>**Quality rating:** Low | **Participants:** 267 housing insecure, pregnant women<br>**Setting:** Brigham Women's Hospital<br>**Dates of data collection:** 2017-2019 compared with data from 2020 (during the COVID-19 pandemic). | **Primary Findings:**<br>Compares women who enrolled in Bridges to Moms (BTM) from 2017-2019 (n=134) with a matched group of housing-insecure women who received prenatal care delivered at BWH from 2017- 2019 (n=133).<br>The BTM group had significantly greater postpartum clinic attendance rates and connections to primary care.<br>Women enrolled in BTM for over 30 days pre-delivery (n=92) had significantly higher prenatal clinic attendance rates and their infants who required NICU care had significantly shorter NICU stays than the BWH group.<br>Dose effect underscores the importance of the timing of intervention in maximising health returns.<br>BTM spent 309% more on addressing food and transportation insecurity.<br>With a nimble adoption of telehealth, BTM documented 13% more clinical encounters in 2020, allowing BTM patients to continue to benefit from equitable consistent care during a time of great disparity. | This cohort study from the USA demonstrates that in tandem with traditional obstetrical services, a program that provides community-based case management, nursing, and social support improves maternal and newborn outcomes and is resilient under pandemic pressures. |
| **Chatzifotiou and Andreadou 2021**<br>**Greece**<br>(Chatzifotiou & Andreadou, 2021) | **Study Design:** Qualitative study investigating the impact of COVID-19 on domestic abuse in Greece.<br>**Data collection methods:**<br>Interviews were carried out at a place and time convenient to the participants | **Participants:**<br>Participants (n=15) came from the northern part of Greece, still live with their abusive partners, and have asked for help and support from formal services in their communities.<br>All participants had experienced physical, verbal, psychological | **Primary Findings:**<br>Important that social welfare professionals (SWPs), such as psychologists and social workers, who treat women victims of IPV, are properly trained in empowerment and coping-focused approaches and are gender-sensitive to confidently deal with the complexity of IPV. | During Wave 1 of the COVID-19 pandemic in March-April 2020, women in northern Greece reported pre-existing and new domestic threats including withdrawal of hand washing rights, and withdrawal of freedoms to use personal protective equipment to prevent COVID-19 infection. |
|  | Each interview lasted, on average, one hour.<br>Interviews were conducted in person by one of the researchers during the lockdown period, considering and following all necessary safety and protective measures.<br>**Quality rating:** Moderate | abuse, and some had experienced intimate partner violence (IPV).<br>Ages of the participants ranged from 30 to 50 years.<br>**Setting:** Interviews were conducted in person by one of the researchers during the lockdown period, considering and following all necessary safety and protective measures.<br>**Dates of data collection:** Semi-structured interviews were conducted during the COVID-19 lockdown of March and April 2020. | SWPs should consider the specific sociocultural context of the women clients.<br>The Greek culture and society, with its patriarchal structured institutions and unequal gender rights, play an important role in the ways that SWPs empower and support women clients.<br>**Additional Findings:**<br>Many participants discussed how the number of threats (a kind of psychological abuse) increased during the COVID-19 lockdown. Participant 13 said, *“He threatened ... that if I coughed, he would drive me out of the house along with the children and ... told me that if we got infected it would be my fault ... he would not let us wash our hands ... nor use antiseptics. I was afraid that my children would catch coronavirus”*. |  |
| **Kossek et al 2021**<br>**USA** | **Study Design:** Qualitative study to understand how women working in Science, Technology Engineering and Mathematics (STEM) varied in adapting to boundary disruptions to manage their work and non-work roles.<br>**Data collection methods:**<br>Participants completed an online survey to acquire qualitative data.<br>**Quality rating:** Moderate | **Sample size:** 763<br>**Participants:** Tenure-stream (on the path to professorship) academic women in STEM and related fields from 202 universities nationwide.<br>**Setting:** The survey was shared on different STEM online mailing lists to obtain a representative cross-section of STEM disciplines.<br>**Dates of data collection:** October 2020 | **Primary Findings:**<br>Care responsibility and family management is often provided by women.<br>Most participants had child or elderly family member responsibilities (72.5%).<br>Many women adapt their way of manoeuvring as a result of these obligations.<br>Women often conceal their non-work roles or reveal them with the hope of changing the working norms of ‘professional image management’. | Results from this quantitative study highlight the difficult decisions faced by STEM women when the scaffolding support network around them was no longer available, and how this lack of a support network influenced the work and non-work interface. |
|  |  |  | • Some women sacrifice roles, whether in the workplace or outside of the workplace.<br>• Social support lessens the effects of professional image management.<br>• Structural support (i.e., tenure-stream extensions) helps STEM women manage their responsibilities.<br>• Allowing STEM women flexibility in where they work benefits mothers, depending on where they can be most productive. |  |
| **Moen et al 2021**<br>**France/ Online** | **Study Design:** Qualitative study of peer mentoring to ensure diversity in health informatics, to target systemic inequalities and build sustainable, intergenerational communities, improve digital health literacy, and build capacity in digital health without losing the human touch.<br>**Data collection methods:**<br>• An online document co-authored by audience and speakers from a MedInfo2019 workshop highlighted the main issues in diversity in health informatics.<br>• This document laid the foundation for all authors to attend an online MIE2020 panel to discuss strategies to | **Participants:** Audience and speakers of the MedInfo2019 conference<br>**Setting:** The initial document was created in Lyon, France at MedInfo2019 and the strategies from MIE2020 were devised online.<br>**Dates of data collection:** MedInfo2019 occurred 25<sup>th</sup> -30<sup>th</sup> August 2019, and MIE2020 occurred 28<sup>th</sup> April to the 1<sup>st</sup> May 2020 | **Primary Findings:**<br>• Medtronic are taking bold steps towards elevating women in a holistic approach building an inclusive, diverse and equitable workspace for women, by providing a variety of development programs for managers, including unconscious bias training and facilitating networks of women and men that promote change (15000 members, 118 hubs, 68 countries).<br>• The McKinsey Global Institute (MGI) has estimated that advancing gender equality could add \$12 trillion per year to the world economy by 2025.<br>• In their study of 90 entities and 50,000 managers, MGI found that companies with more women in executive teams had a 56% higher operating profit, highlighting that gender-balanced organisation produced results that were | • This qualitative paper from France does not include a scientific study design.<br>• Strategies were organised from conference panel discussions.<br>• Strategies currently in place have not been evaluated. |
|  | ensure diversity in health informatics.<br>**Quality rating:** Low |  | far more predictable, sustainable, and profitable.<br>• The UN Sustainable goals targets gender equality (SDG 5), reducing inequality (SDG 10), and cross-sectional collaborative actions (SDG 17). |  |

**Abbreviations:** **2S people** - Two spirit people [an important term within some Indigenous cultures and some Indigenous people, meaning a person with both a feminine and a masculine spirit living in the same body]. **ARIMA** - Autoregressive integrated moving average. **BTM** - Bridges to Moms. **CIHR** Canadian Institutes of Health Research **PI** – Principal Investigator **SDG** – [UN] Sustainable Development Goals **STEM** – Science, Technology, Engineering, and Mathematics.

**Quantitative studies:** Five (n = 5) primary papers were quantitative studies (Davic et al., 2021; Millán et al., 2021; Ray et al., 2021; Witteman et al., 2021; Zhang et al., 2021)

**Qualitative studies:** Three (n = 3) primary papers were qualitative studies (Chatzifotiou & Andreadou, 2021; Kossek et al., 2021; Moen et al., 2021)

### 2.3 Work domain

The six included work domain papers were comprised of one review (Richey & Pointer, 2021); one quantitative survey (Millán et al., 2021); one cohort study (Witteman et al., 2021), two qualitative studies (Kossek et al., 2021; Moen et al., 2021) and one commentary paper (Banks et al., 2021).

The commentary (Richey & Pointer, 2021) reported a case study from the USA investigating cognitive behavioural therapy (CBT) as a therapeutic modality to use with women healthcare workers in response to the pervasive stressors of the COVID-19 pandemic. The authors stated that the CBT modality requires further empirical testing to see how it could be integrated in wider settings with women of colour to provide an ‘anti-racist intersectional framework’.

The quantitative study (Millán et al., 2021) surveyed labour market trends in Spain over a 12 year period which included the first year of the pandemic. The authors advocated for more stable jobs for women to increase the percentage of permanent contracts and complete working days. Permanent contracts and full-time hours would increase income for women, and thereby decrease the existing wage gap that is not exclusive to Spain (Millán et al., 2021).

Conducted in Canada, a cohort study (Witteman et al., 2021) evaluated the investigator-initiated programs of the Canadian Institutes of Health Research (CIHR). The study found that targeting funding calls encouraged female researchers to submit more applications and increased female-led projects from approximately 30% to 40%.

One qualitative study from France (Moen et al., 2021) highlighted peer-mentoring strategies to ensure diversity in health informatics. Another qualitative study from the USA (Kossek et al., 2021) investigated the difficult decisions faced by women working in the areas of Science, Technology, Engineering and Mathematics (STEM) when previous support networks were unavailable during COVID-19 lockdowns. The work and non-work interface changed immensely with increased stress related to balancing responsibilities at work and within the home. Qualitative interviews regarding new ways of working and disrupted boundaries included the following quote:

> “I am saying ‘no’ more often to work opportunities or responsibilities. Also. I am sometimes now saying ‘no’ to my family and making them work it out for themselves. The latter has not always been a good thing short term, I can’t tell long term. I am also not planning fun activities for us as a family or planning vacations or events. I am not helping others in my family as much to work out differences. Maybe this will have some positive long-term effects, but short term we are less connected as a family, and my husband and I are significantly less happy”. (Within the theme of work contextual features associated with adapting to disrupted boundaries). Page 1620 (Kossek et al., 2021).

One commentary paper from Canada (Banks et al., 2021) discussed several strategies to reduce the effects of gender inequality for women. These included:

- Funding a national childcare program to increase women’s income and participation in the workforce (Canadian Government)
- Improving mentoring and sponsorship of women and marginalised individuals (Canadian Cardiovascular Society Equity and Diversity Initiatives)
- Implementing sex and gender training for researchers submitting grants (Canadian Institutes for Health Research)
- Targeting awards for sex- and gender-based analyses (Canadian Institutes for Health Research)
- Ensuring equal representation, including women and marginalised persons, in pandemic response and recovery planning and decision-making

**Bottom line for work domain: Permanent contracts, full-time hours, national childcare programmes, and peer mentoring could increase employment and income for women, and thereby could help to decrease the existing wage gap.**

### 2.4 Health domain

The six included health domain papers included one systematic review (Saad et al., 2021); one rapid review (Banati & Idele, 2021), one rapid scoping review (Steinert et al., 2021); one concept paper (Das et al., 2021); one viewpoint paper (Figueroa et al., 2021); and one quantitative survey (Ray et al., 2021).

A systematic review (Saad et al., 2021) conducted in Canada found that COVID-19 had increased barriers to accessing mental health care for pregnant and postpartum women who were experiencing increased and worsened levels of depression and anxiety.

A rapid review (Banati & Idele, 2021) suggested that interventions such as helplines, virtual safe spaces and online counselling may help address issues of violence and abuse experienced by women and girls during COVID-19 lockdowns and restrictions. This increase in violence and abuse has negatively affected the mental health of women and girls who have also assumed increased caring responsibilities.

A rapid scoping review (Steinert et al., 2021) examined evidence from several countries relating to different pandemics (COVID-19, Ebola and Zika). The review found that online or telephone interventions could be appropriate in supporting women. For example, during the Zika outbreak in Puerto Rico, social media platforms such as Facebook provided a helpful tool for distributing reproductive health messages.

A concept paper (Das et al., 2021) from the USA suggested that professional women were more likely to be overlooked in terms of leadership and visibility. During the pandemic, academics and leaders frequently reverted to working with ‘who they know’ which meant that women were often less visible in leadership roles.

A viewpoint paper (Figueroa et al., 2021) from the USA illustrated gender inequality issues in digital health at three levels: users, tools (i.e., mobile phone applications) and leadership.

The paper offered recommendations for tackling gender bias through a feminist intersectional framework. For example, leadership in digital health companies was more likely to be male, leading to exclusion of women from developing technology to target women’s health.

A quantitative survey (Ray et al., 2021) of university students in the USA during the pandemic found that women reported greater levels of COVID-19 related worry and anxiety than men. Women also experienced a greater number of daily life disruptions. Women experienced more difficulty in sleeping, greater loss of income and increased reduction in work hours (Ray et al., 2021).

**Bottom line for health domain: Interventions such as helplines, virtual safe spaces, and online counselling could help improve the mental wellbeing of women and girls. Digital health companies could benefit from employing women executives to develop technology targeting women’s health.**

### 2.5 Living standards domain

The six included living standards papers included four reviews (Andermann et al., 2021; Bray & McLemore, 2021; Grammatikopoulou et al., 2021; Perri et al., 2021), one cohort study (Zhang et al., 2021); and one case study (Goodsmith et al., 2021).

The four review papers were comprised of a narrative review conducted in Greece (Grammatikopoulou et al., 2021); a scoping review conducted in Canada (Andermann et al., 2021), a scoping review conducted in the USA (Bray & McLemore, 2021), and a literature review conducted in the USA (Perri et al., 2021).

The narrative review (Grammatikopoulou et al., 2021) indicated that care provided to women experiencing homelessness during the pandemic should be optimised at multidimensional levels. This includes ensuring access to safe water and sanitation, quality food, psychological support, disease management, acute health care, opportunities for employment, and support for minor dependents.

The scoping review from Canada (Andermann et al., 2021) indicated that a ‘population approach’ could improve outcomes for women and their children. A population approach involves investigating the causes of homelessness among women, such as intimate partner violence. To avoid crisis situations, such as homelessness, a number of structural changes were recommended to promote gender equality in living standards:

- Formalising care work with pay scales and benefits
- Improving access to childcare
- Ensuring pay equity
- Creating opportunities for parental leave and work-life balance
- Helping support families in creating stronger adult-adult and adult-child attachments
- Nurturing social-emotional competencies for families through prenatal classes
- Creating greater family stability by reducing intimate partner violence and adverse childhood experiences, which are often precursors to homelessness.

Evidence from a USA scoping review (Bray & McLemore, 2021) identified scientific and structural racism leading to gaps in data that fail to account for the realities of black life, including the maternal health of black women. Several action steps were suggested:

- Targeting investments for black students, educators, healthcare providers, and researchers
- Putting the voices, strategies, and interventions of black birthing people at the centre of decisions about maternal health
- Establish authentic partnership with black women and female-led organisations to reduce maternal morbidity and mortality (Bray & McLemore, 2021).

A literature review from the USA (Perri et al., 2021) described the implications of a lack of gender redistributive/transformative approaches in sectors, such as housing and employment, that impact living standards. In the employment sector, for example, sexual and gender minorities (SGM) were more likely to lose employment (17%) during the pandemic compared with the general population (13%). SGM were also much less likely to have access to paid sick leave (29%) compared with the general population (76%).

A cohort study from the USA (Zhang et al., 2021) demonstrated that community-based case management, nursing, and social support for homeless women improved maternal and new born outcomes. Homeless women enrolled on the Bridges to Moms (BTM) programme for over 30 days pre-delivery (n = 92) reported significantly higher prenatal clinic attendance rates and significantly shorter neonatal intensive care unit stays than a comparison group.

A case report from the USA (Goodsmith et al., 2021) focused on homeless pregnant women who were survivors of domestic violence. Cross-agency collaboration provided these women with safe interim housing. Examples include Project Roomkey, which made several thousand hotel rooms available to accommodate vulnerable women with high-risk medical conditions during the pandemic, and Project Safe Haven which provided safe and adequate accommodation in hotel rooms for approximately 900 survivors of domestic violence and their children.

**Bottom line for living standards domain: Care provided to women experiencing homelessness could be optimised at a multidimensional level. Interventions such as Project Roomkey and Safe Haven are reported examples of providing homeless women and their children with safe and adequate accommodation in hotels.**

### 2.6 Personal security domain

Two studies focused on personal security. One primary study from Greece (Chatzifotiou & Andreadou, 2021) found that the experience of being in ‘lockdown’ with an abusive partner during the pandemic created threats to well-being and life. Specific training of social workers, psychologists and therapists is needed to empower women to use coping strategies and utilise services to get away from abusive partners.

A commentary paper from the USA (Hinton et al., 2021) suggested several ways in which multiple sectors can work together to ensure adequate support for female survivors of domestic violence:

- Reinforcing and extending emergency phones and 24-hour hotlines, and temporary shelters for survivors.
- Equipping first responders of violence against women to make prompt referrals to support services.
- Promptly processing complaints and protection orders.
- Adjusting lockdown restrictions during a pandemic (e.g., in Spain, women who leave a situation of domestic violence are exempt from lockdown)
- Expanding technology-based solutions (e.g., smartphone applications that could be used during lockdown restrictions to increase access to information on violence against women, service provision, and data collection.

**Bottom line for personal security domain: Specific training of social workers, psychologists, and therapists to empower women to utilise services to get away from abusive partners during times of lockdown are needed. Multiple sectors working together to provide 24-hour hotlines, temporary shelters, prompt referrals to support services, and protection orders are needed.**

### 2.7 Participation domain

Only one cross-sectional study (Davic et al., 2021) from the USA investigated gender representation of academic conference presenters. The study reported that gender disparities in the representation of academic/professional work could be reduced by more frequent use of online platforms. Women with caring and childbearing responsibilities were more likely to attend and present at conferences when they were held online. Women with childcare responsibilities are less likely to attend face to face conferences which involve more time away from home.

**Bottom line for participation domain: Gender disparities in the presentation of academic/professional work could be reduced by more frequent use of online conferences.**

### 2.8 Education domain

There were no peer-reviewed papers identified in the search within the domain of education. Two reports in the grey literature described interventions that focused on girls’ education, but no evaluation data was provided (Gender Equality Advisory Council, 2021; Hammond, Matulevich, Beegle, & Kumaraswamy, 2020).

A G7 intergovernmental forum report (Gender Equality Advisory Council, 2021) made several recommendations for gender-transformative education, including supporting girl-led groups and activists by ensuring accessible information and providing flexible funding. In order to eliminate stereotypes and unconscious bias at all levels of education, teacher training curricula should empower teachers to understand and challenge gender stereotypes in learning environments.

A report from the Gender Group at the World Bank Group (Gender Equality Advisory Council, 2021) focused on advancing the participation of women and girls in STEM. It highlighted Technovision, a successful entrepreneurship programme for girls aged 10-18.

Working with female mentors, girls identified a community problem and then developed a mobile application to help solve the problem. Some of the applications developed by more than 30,000 girls who have completed the programme have addressed such problems as food waste, lack of nutrition and women’s safety issues.

The World Bank has also developed Teach, a free classroom observation tool with a component to measure the extent to which the teacher challenges gender bias in the classroom. The World Bank is also developing Coach, which involves training materials for teachers to address gender stereotypes and biases in the classroom.

**Bottom line for education domain: There is a lack of research in this area. Teacher training curricula could empower teachers to understand and challenge gender stereotypes in learning environments. Education for girls should enable participation in STEM, as exemplified in the Technovision programme.**

## 3. DISCUSSION

### 3.1 Summary of the findings

A total of 21 peer-reviewed papers were included in this rapid review: 7 were reviews, 6 were commentaries, and 8 were primary studies. The included papers were related to the six EHRC domains: work, health, living standards, personal security, participation, and education. Work

The COVID-19 pandemic created challenges for many women working from home while providing elderly care, family care and childcare simultaneously (Kossek et al., 2021; Moen et al., 2021; Richey & Pointer, 2021). These challenges often disrupted work and home-life in negative ways, reducing work confidence and productivity and potentially increasing the gender gap in wages. COVID-19 lockdowns and restrictions resulted in more men moving forward in their careers while women faltered or slowed down. Research suggests that there should be more emphasis on creating permanent full-time jobs for women to reduce the gender gap in wages (Millán et al., 2021). In Canada, targeting female research leaders was a successful strategy in reducing the gender gap of securing research funding in Canada (Witteman et al., 2021).

### 3.2 Health

Online platforms, virtual spaces, digital health and technology can aid women in mental wellbeing, reproductive health and mitigating gender-based violence (Banati & Idele, 2021; Das et al., 2021; Ray et al., 2021; Saad et al., 2021; Steinert et al., 2021). Digital health was especially effective during the COVID-19 pandemic when social distancing and lockdown measures were operationalised (Banati & Idele, 2021). Digital health can also deliver important health messages in accessible online formats, such as via Facebook (Steinert et al., 2021). Future digital initiatives targeting women’s health will benefit from women being actively involved in the design and the development of gender-friendly technology that meets the needs of women (Figueroa et al., 2021).

### 3.3 Living standards

Women, racialised people, immigrants, people with disabilities, sexual and gender minorities and people at the intersections of those groups are more likely to experience unemployment and underemployment, associated occupational safety risks, and poorer health leading to lower standards of living (Perri et al., 2021).

The Bridges to Moms project provides an antenatal outreach programme for homeless mothers, thus preventing long stays in neonatal intensive care units (Zhang et al., 2021). The importance of providing multidimensional care to homeless women was emphasised (Grammatikopoulou et al., 2021). This care includes adequate water and sanitation, quality food, psychological support, disease management, acute health care, opportunities for employment and support for minor dependents. Project Roomkey and Safe Haven were two interventions that provided safe accommodation for homeless mothers and their children during the pandemic (Goodsmith et al., 2021).

### 3.4 Personal security

To protect women from abusive partners, health and social care professionals (i.e., social workers, psychologists, therapists) could benefit from specialised training to empower women in coping strategies and utilising available services (Chatzifotiou & Andreadou, 2021). In addition, smartphone applications can make it easier for women to access services and document abuse in a safe, secure, and legally admissible way (Hinton et al., 2021).

### 3.5 Participation

Gender disparities in the representation of academic/professional work could be reduced by more frequent use of on-line platforms. Women with caring and childbearing responsibilities are more likely to attend and present at online conferences. Face to face conferences involve more time away from home due to travelling (Davic et al., 2021).

### 3.6 Education

To eliminate stereotypes and unconscious bias at all levels of education, teacher training curricula should empower teachers to understand and challenge gender stereotypes and gender bias in learning environments (Gender Equality Advisory Council, 2021). In addition, innovations to advance the participation of girls in STEM should be supported, such as Technovision, in which girls work with female mentors to identify a community problem and then develop a mobile application to solve the problem.

### 3.7 Cost-effectiveness

No published studies have been reported on the costs or cost-effectiveness of innovations to lessen gender inequalities due to the COVID-19 pandemic. However, there are explicit costs of failing to carry out equality and diversity innovations/interventions regarding gender equality.

### 3.8 Evidence gaps

There was limited evidence on interventions for reducing gender inequalities in the domains of, personal security and participation, and a lack of research evidence for educational innovations in OECD countries (including Wales). There was a lack of data, disaggregated by race, ethnicity, gender, income and locality. More research is needed on how and why gender inequalities are perpetuated through social infrastructure and what could be done to affect change (Toure, Langlois, Shah, McDougall, & Fogstad, 2021).

### 3.9 Limitations of available evidence

Limitations of this rapid review include a lack of high-quality evidence such as RCTs and service evaluation studies that evaluate specific innovations or tailored services to reduce gender inequalities. Only 8 papers in this rapid review were primary studies indicating that more primary research is needed to evaluate specific innovations or tailored interventions to reduce gender inequalities in the UK context.

### 3.10 Implications for policy and practice

Implications for policy and practice are outlined below for each of the six EHRC domains:

#### 3.10.1 Work

- Interventions/innovations/policies related to work include:

- Permanent contracts, full-time hours, and national childcare programmes are recommended to increase income for women and thereby decrease the existing gender wage gap.
- Prioritising accessibility of affordable childcare through increased public investment.
- Introduce gender equality criteria in public sector procurement and targets for public spending on women-owned and women-led businesses.
- More frequent use of online platforms in the presentation of professional work can reduce gender disparities due to time saved in travel away from home.

#### 3.10.2 Health

- Interventions/innovations/policies related to health include:

- Access to antenatal health support for homeless or vulnerable mothers should remain available even when social distancing is needed.
- Digital health information should be available to women on accessible online or social media platforms.
- Leadership in digital health companies could benefit from women developing gender-friendly technology that meets the health needs of women.
- Authentic partnerships with black women and female-led organisations should be developed to reduce maternal morbidity and mortality.

#### 3.10.3 Living Standards

- Interventions/innovations/policies related to living standards include:

- Multi-dimensional care is needed for women experiencing homelessness.
- Community-based nursing care and social support for homeless women is needed to improve maternal and new-born outcomes.
- Interventions such as Project Roomkey and Safe Haven (Goodsmith et al., 2021) are examples of providing homeless women and their children with safe hotel accommodation.

#### 3.10.4 Personal Security

- Interventions/innovations/policies related to personal security include:

- Specific training of social workers, psychologists and therapists is needed to empower women to use coping strategies and utilise services to gain protection from abusive partners.
- Interventions such as helplines, virtual safe spaces and online counselling could help address violence and abuse experienced by women and girls.
- Smartphone applications could be used to make it easier for women to access services and document abuse in a safe, secure and legal way.

#### 3.10.5 Participation

- Interventions/innovations/policies related to participation include:

- Frequent use of online platforms can reduce gender disparities in the representation in academic/professional work.
- Conference organisers should consider the gender of speakers and keynote presenters within conferences.
- Equal representation, including women and marginalised persons, is important in planning pandemic recovery programmes

#### 3.10.6 Education

- An evidence gap was identified in this area; innovations identified are from the grey literature with a lack of intervention/innovation evaluation data. These include:

- Teacher training curricula to empower teachers to understand and challenge gender stereotypes in learning environments.
- Education for girls to enable participation in STEM.

#### 3.10.7 Further research

Innovations implemented in the domains of personal security, participation and education require robust evaluation.

Further research is required to understand the effectiveness of gender equality innovations for minority groups.

### 3.11 Strengths and limitations

#### 3.11.1 Strengths

This rapid review focused on the peer-reviewed papers (n=21) in alignment with the six domains identified by EHRC. The 21 peer-reviewed papers were a mixture of reviews, commentaries, and primary studies. Grey literature (n=14) is included in Appendix 2. The rapid review investigated innovations/interventions to reduce gender inequality during the first three waves of the COVID-19 pandemic between March 2020 and December 2021. Data was not presented according to ‘waves’, but data collection dates are provided in the data extraction tables in Appendix 1.

#### 3.11.2 Limitations

Due to various study designs, a quality appraisal was conducted with a variety of checklist tools, making comparisons difficult between studies. Although efforts were made to include education studies, no peer-reviewed studies were found for this domain. No economic evaluation studies were found to investigate the cost-effectiveness of innovations to reduce gender inequalities during the COVID-19 pandemic. A narrative synthesis was used to describe the studies.

## Data Availability

All data produced in the present study are available upon reasonable request to the authors

## 5. RAPID REVIEW METHODS

### 5.1 Literature search

The PICO and eligibility criteria are presented in Table 1. Evidence sources and the search strategy for this rapid review is presented below.

### 5.2 Evidence sources

Key evidence sources included:

1. Medline
2. CINAHL
3. PsycInfo
4. Cochrane Library
5. ASSIA
6. EmBASE

Key sources were searched for papers published between 1 January 2020 and 13 December 2021. The searches were limited to published research in the English language. The scope outlined for this search was to keep the review concise and deliverable within the timeframe expected for a rapid review.

### 5.2 Search strategy

Below is an example of a search strategy for the Medline database.

#### Search strategy (Medline)

1. exp Gender Equity/
2. exp Sexism/
3. (equal* or equalit* or inequalit* or inequit* or equit* or discriminat* or sexism or disadvantage).ti,ab.
4. 1 or 2 or 3
5. exp Women/
6. (female* or feminin* or woman or women or transwomen or girl*).ti,ab.
7. 5 or 6
8. exp Coronavirus/
9. exp COVID-19/
10. ((corona* or corono*) adj1 (virus* or viral* or virinae*)).ti,ab,kw.
11. (coronavirus* or coronovirus* or coronaviri* or 2019-nCoV or 2019nCoV or nCoV2019 or nCoV-2019 or COVID-19* or COVID19* or ncov* or n-cov* or HCoV* or SARS-CoV-2 or SARSCoV-2 or SARSCov2 or SARS-CoV2 or severe acute respiratory syndrome).ti,ab,kw.
12. ((outbreak* or pandemic* or epidemic*) adj10 (wuhan or hubei or china or Chinese or Huanan)).ti,ab,kw.
13. 8 or 9 or 10 or 11 or 12
14. 4 and 7 and 13

See Table 5 for the names of the databases searched including number of papers found in each database.

**Table 5:**
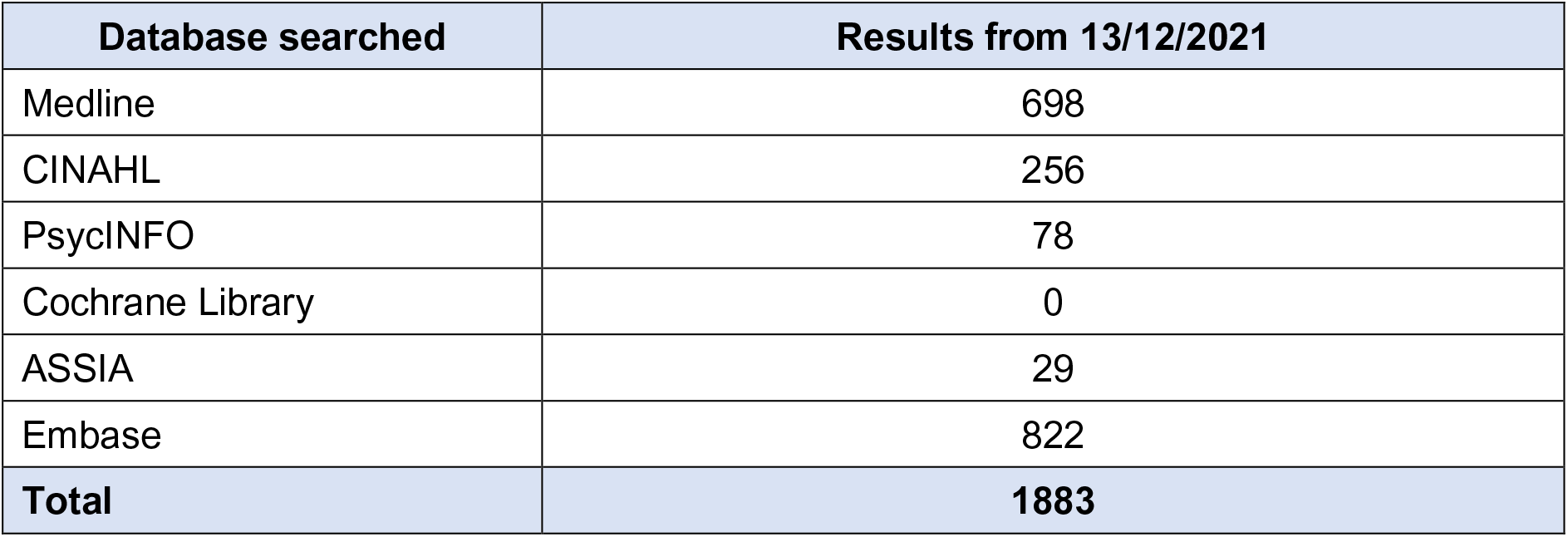
Databases searched.

### 5.3 Reference management

The COVIDence reference management system was used to store and manage citations. Duplicates were removed in COVIDence (Veritas Health Innovation, 2021).

### 5.4 Study selection process

Using the COVIDence tool, citations were screened on title and abstract by members of the review team. Full-text articles were then retrieved and further assessed for inclusion. Any queries regarding inclusion/ exclusion were resolved by discussion between members of the review team.

Due to the time constraints of a rapid review, full double screening was not possible. However, a sample of citations were double screened by the review lead to ensure adherence to inclusion/exclusion criteria.

### 5.5 Data extraction

The data was extracted from the included studies using a pre-defined data extraction tool to capture all relevant data. Extracted data included study details such as author, year, setting, aim, design, population, and sample size. The data extraction also included data specific to the review question, type of study, method of analysis, key findings, and author conclusions.

Included papers were distributed among the review team for data extraction. A sample of extracted studies was checked against the papers for accuracy by the review lead. A proportion of the papers (10%) were double extracted to check for discrepancies between reviewers.

### 5.6 Quality appraisal

Quality appraisal was conducted by members of the review team using the JBI critical appraisal tools which include the JBI analytical cross-sectional study checklist (Joanna Briggs Institute, 2017a), JBI systematic reviews and research syntheses checklist (Joanna Briggs Institute, 2017c), JBI case reports checklist (Munn et al., 2021), and JBI cohort studies checklist (Joanna Briggs Institute, 2021).

Members of the review team chose the most appropriate JBI critical appraisal tool. A quarter of critical appraisals will be checked by a second reviewer. Discrepancies arising during the critical appraisal process were discussed until an agreement was reached by the review team.

## 6. EVIDENCE

### 6.1 Study selection flow chart

The study selection flow chart is shown in Figure 1 as a PRISMA flow chart (Page et al., 2021).

**Figure 1.**
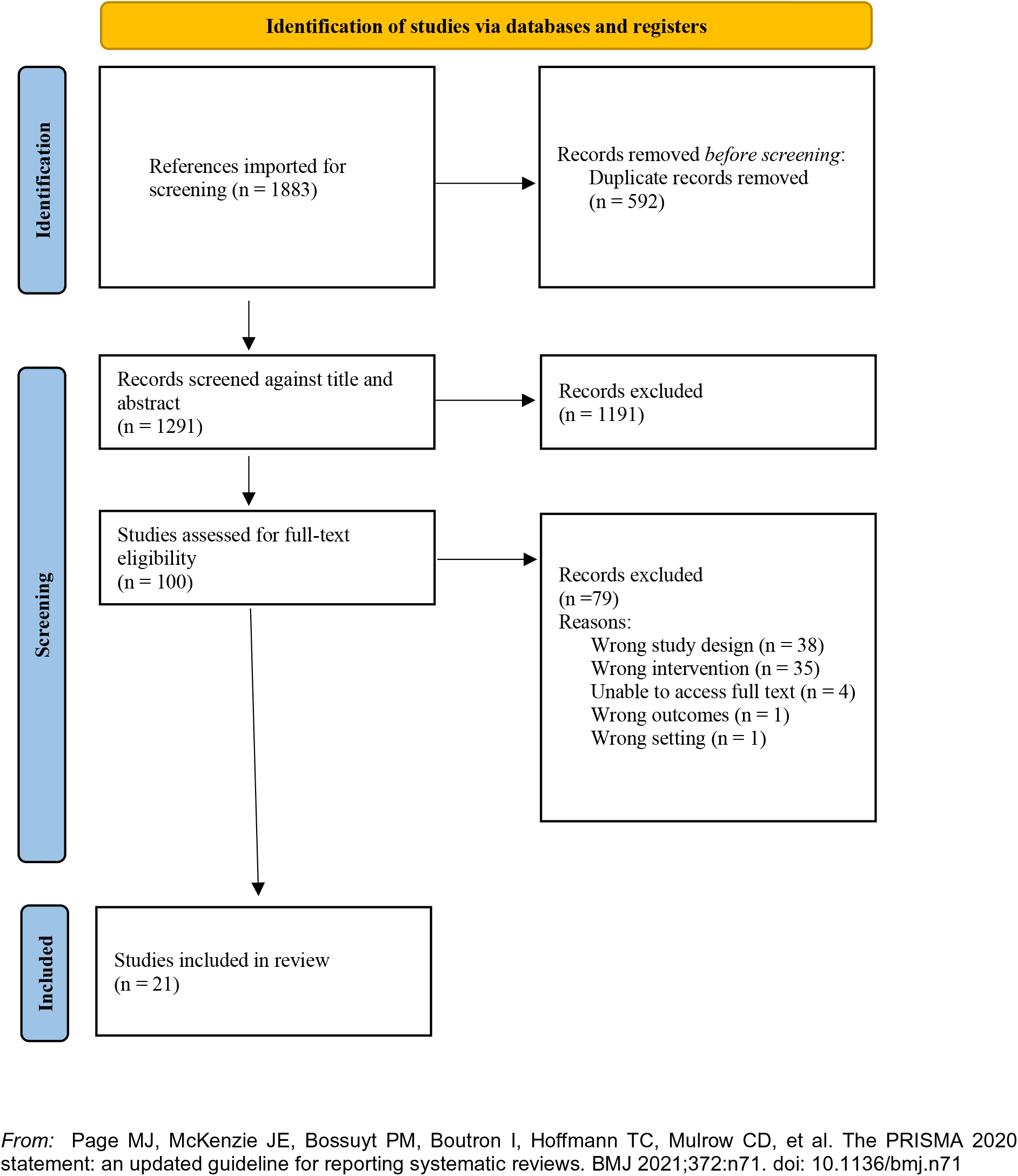
PRISMA study selection flowchart (Page et al., 2021)

### 6.2 Data extraction tables

The data extraction tables are presented in the results section (see Tables 2, 3 and 4).

## 7. ADDITIONAL INFORMATION

### 7.2 Acknowledgements

The authors would like to thank expert stakeholders from the Equality Inclusion and Human Rights team at Welsh Government, including Hannah Fisher (Senior Policy Advisor), Steven Macey (Senior Research Officer), Laura Price (Gender Equality Review Manager) and Emma Bennett (Head of Equality, Inclusion and Human Rights). Dr Ruth Lewis and Dr Alison Cooper (Wales COVID-19 Evidence Centre) for providing support to the review team; and Elizabeth Gillen, Cardiff University, for reviewing our search strategy and conducting the Embase search for this Rapid Review.

### 7.3 Author contributions

Project leads: CW, RTE, NB, DH; drafting of report: LHS, RTE, NH; contribution to writing and critical editing of the report; LHS, NH, AH, BA, AM, KP, NN, CW, RTE, DH, DF; reviewing: LHS, BA, NH, AH, AM, KP, RG, JD.

### 7.3 Disclaimer

Disclaimer: The views expressed in this publication are those of the authors, not necessarily Health and Care Research Wales. The WCEC and authors of this work declare that they have no conflict of interest.

## 8. ABOUT THE WALES COVID-19 EVIDENCE CENTRE (WCEC)

The WCEC integrates with worldwide efforts to synthesise and mobilise knowledge from research.

We operate with a core team as part of Health and Care Research Wales, are hosted in the [Wales Centre for Primary and Emergency Care Research (PRIME)](http://www.primecentre.wales/), and are led by [Professor Adrian Edwards of Cardiff University](https://www.cardiff.ac.uk/people/view/123022-edwards-adrian).

The core team of the centre works closely with collaborating partners in [Health Technology Wales](https://healthtechnology.wales/), Wales Centre for Evidence-Based Care, [Specialist Unit for Review Evidence centre](https://www.cardiff.ac.uk/specialist-unit-for-review-evidence), [SAIL Databank](https://healthandcareresearchwales.org/about-research-community/secure-anonymised-information-linkage-sail-databank), Bangor Institute for Health & Medical Research/ Health and Care Economics Cymru, and the [Public Health Wales Observatory](https://phw.nhs.wales/services-and-teams/observatory/).

Together we aim to provide around 50 reviews per year, answering the priority questions for policy and practice in Wales as we meet the demands of the pandemic and its impacts.

**Director:**

Professor Adrian Edwards

**Contact Email:**

**Website:**

https://healthandcareresearchwales.org/about-research-community/wales-COVID-19-evidence-centre

## 9. APPENDICES

## Appendix 1: Quality appraisal tables

Members of the review team chose the most appropriate JBI critical appraisal tool. A quarter of critical appraisals will be checked by a second reviewer. Discrepancies arising during the critical appraisal process will be discussed until an agreement is reached by the review team. When possible, studies will be graded as ‘very low’, ‘low’, ‘moderate’ or ‘high’ quality (See Table A1-A7 below).

**Table A1.**
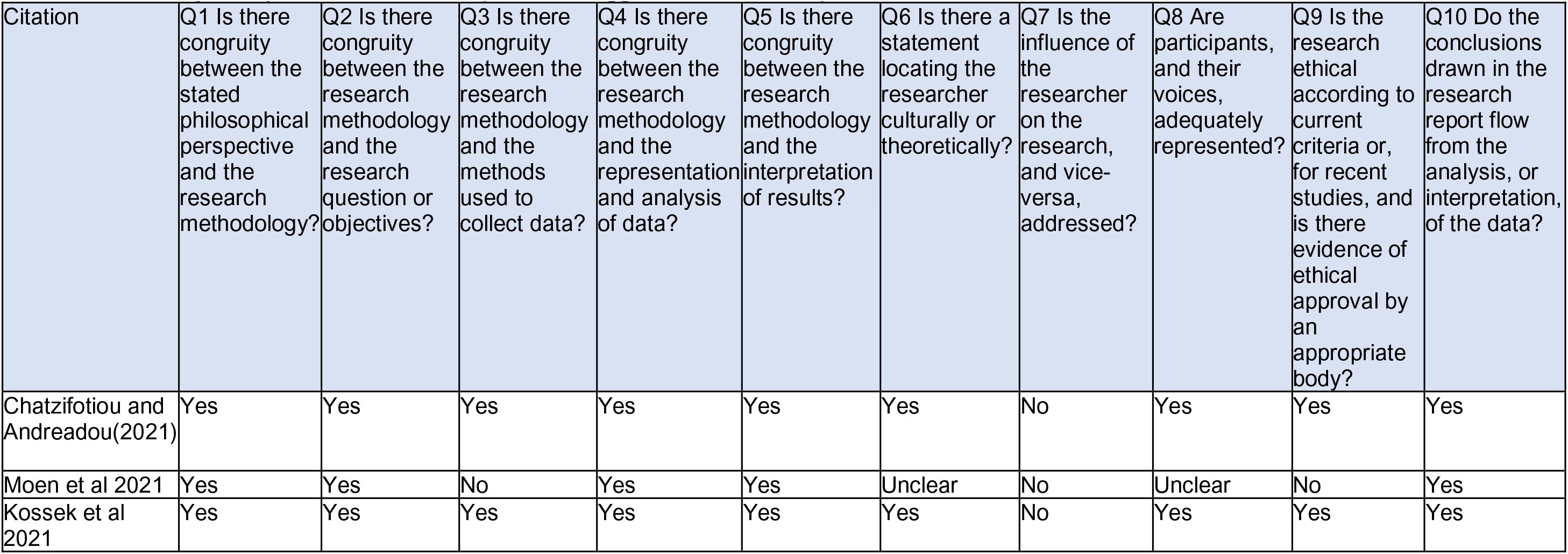
JBI analytical qualitative checklist (Joanna Briggs Institute, 2017b)

**Table A2.** JBI analytical cross-sectional study checklist (Joanna Briggs Institute, 2017a)

| Citation | Q1. Were the criteria for inclusion in the sample clearly defined? | Q2. Were the study subjects and the setting described in detail? | Q3. Was the exposure measured in a valid and reliable way? | Q4. Were objective, standard criteria used for measurement of the condition? | Q5. Were confounding factors identified? | Q6. Were strategies to deal with confounding factors stated? | Q7. Were the outcomes measured in a valid and reliable way? | Q8. Was appropriate statistical analysis used? |
| --- | --- | --- | --- | --- | --- | --- | --- | --- |
| Davic et al 2021 | Yes | Yes | Yes | N/a | N/A | N/A | Yes | Yes |
| Millan et al 2021 | Yes | Yes | Yes | Yes | Yes | Yes | Yes | Yes |

**Table A3.** JBI cohort checklist (Moola et al., 2017)

| Citation | Q1. Were the two groups similar and recruited from the same population? | Q2. Were the exposures measured similarly to assign people to both exposed and unexposed groups? | Q3. Was the exposure measured in a valid and reliable way? | Q4. Were confounding factors identified? | Q5. Were strategies to deal with confounding factors stated? | Q6. Were the groups/ participants free of the outcome at the start of the study (or at the moment of exposure)? | Q7. Were the outcomes measured in a valid and reliable way? | Q8. Was the follow up time reported and sufficient to be long enough for outcomes to occur? | Q9. Was follow up complete, and if not, were the reasons to loss to follow up described and explored? | Q10. Were strategies to address incomplete follow up utilized? | Q11. Was appropriate statistical analysis used? |
| --- | --- | --- | --- | --- | --- | --- | --- | --- | --- | --- | --- |
| Wittema n et al 2020 | Yes | Yes | Yes | Yes | Yes | Yes | Yes | Yes | Yes | Yes | Yes |
| Zhang et al (2021) | Yes | Yes | Unclear | N/A | N/A | Yes | Yes | Unclear | Yes | Unclear | Unclear |

**Table A4.**
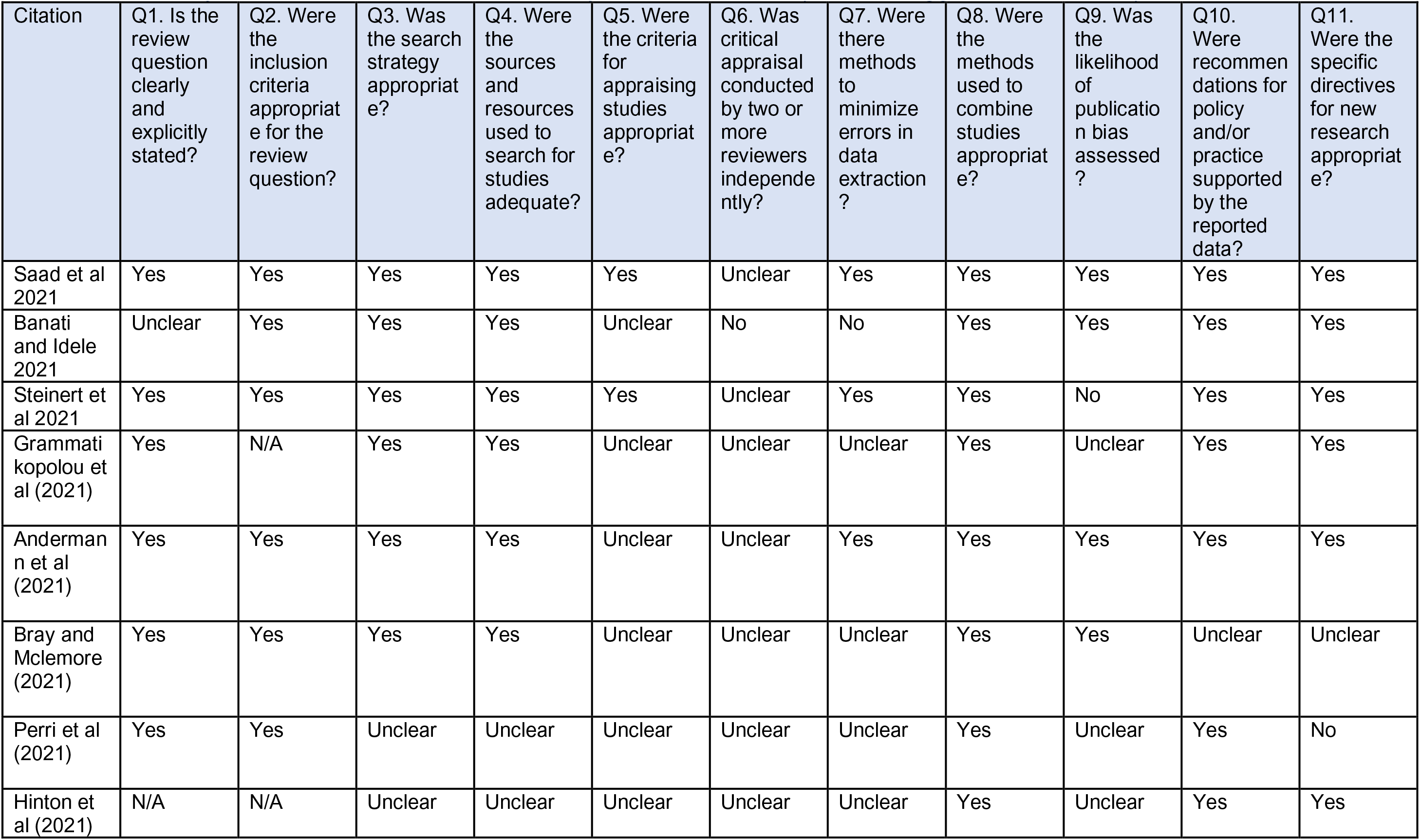
JBI Systematic Reviews and Research Syntheses Checklist (Joanna Briggs Institute, 2017c)

**Table A5.** CEBM Survey quality appraisal checklist (Center for Evidence Based Management, 2005)

| Citation | Q1 Did the study address a clearly focused question / issue? | Q2 Is the research method (study design) appropriate for answering the research question? | Q3 Is the method of selection of the subjects (employees, teams, divisions, organizations) clearly described? | Q4 Could the way the sample was obtained introduce (selection) bias? | Q5 Was the sample of subjects representative with regard to the population to which the findings will be referred? | Q6 Was the sample size based on pre-study considerations of statistical power? | Q7 Was a satisfactory response rate achieved? | Q8 Are the measurements (questionnaires) likely to be valid and reliable? | Q9 Was the statistical significance assessed? | Q10 Are confidence intervals given for the main results? | Q11 Could there be confounding factors that haven't been accounted for? | Q12 12. Can the results be applied to your organization? |
| --- | --- | --- | --- | --- | --- | --- | --- | --- | --- | --- | --- | --- |
| Ray et al 2021 | Yes | Yes | Yes | No | Yes | No | Yes | Yes | Yes | No | Unclear | N/A |

**Table A6.** JBI Critical Appraisal Checklist for Case Reports (Gagnier et al., 2013)

| Citation | Q1. Were patient's demographic characteristics clearly described? | Q2. Was the patient's history clearly described and presented as a timeline? | Q3. Was the current clinical condition of the patient on presentation clearly described? | Q4. Were diagnostic tests or assessment methods and the results clearly described? | Q5. Was the intervention(s) or treatment procedure(s) clearly described? | Q6. Was the post-intervention clinical condition clearly described? | Q7. Were adverse events (harms) or unanticipated events identified and described? | Q8. Does the case report provide takeaway lessons? |
| --- | --- | --- | --- | --- | --- | --- | --- | --- |
| Richey and Pointer 2021 | Yes | Yes | Yes | Yes | Yes | Yes | No | Yes |

**Table A7.** JBI Critical Appraisal Checklist for Case Series (Munn et al., 2021)

|  | Q1. Were there clear criteria for inclusion in the case series? | Q2. Was the condition measured in a standard, reliable way for all participants included in the case series? | Q3. Were valid methods used for identification of the condition for all participants included in the case series? | Q4. Did the case series have consecutive inclusion of the case series? | Q5. Did the case series have complete inclusion of participants? | Q6. Was there clear reporting of the demographics of the participants in the study? | Q7. Was there clear reporting of clinical information of the participants? | Q8. Were the outcomes or follow up results of cases clearly reported? | Q9. Was there clear reporting of the presenting sites(s) / clinic(s) demographic information? | Q10. Was statistical analysis appropriate? |
| --- | --- | --- | --- | --- | --- | --- | --- | --- | --- | --- |
| Goodsmith et al (2021) | Yes | N/A | N/A | N/A | N/A | Yes | Yes | Yes | N/A | N/A |

## Appendix 2: Grey literature

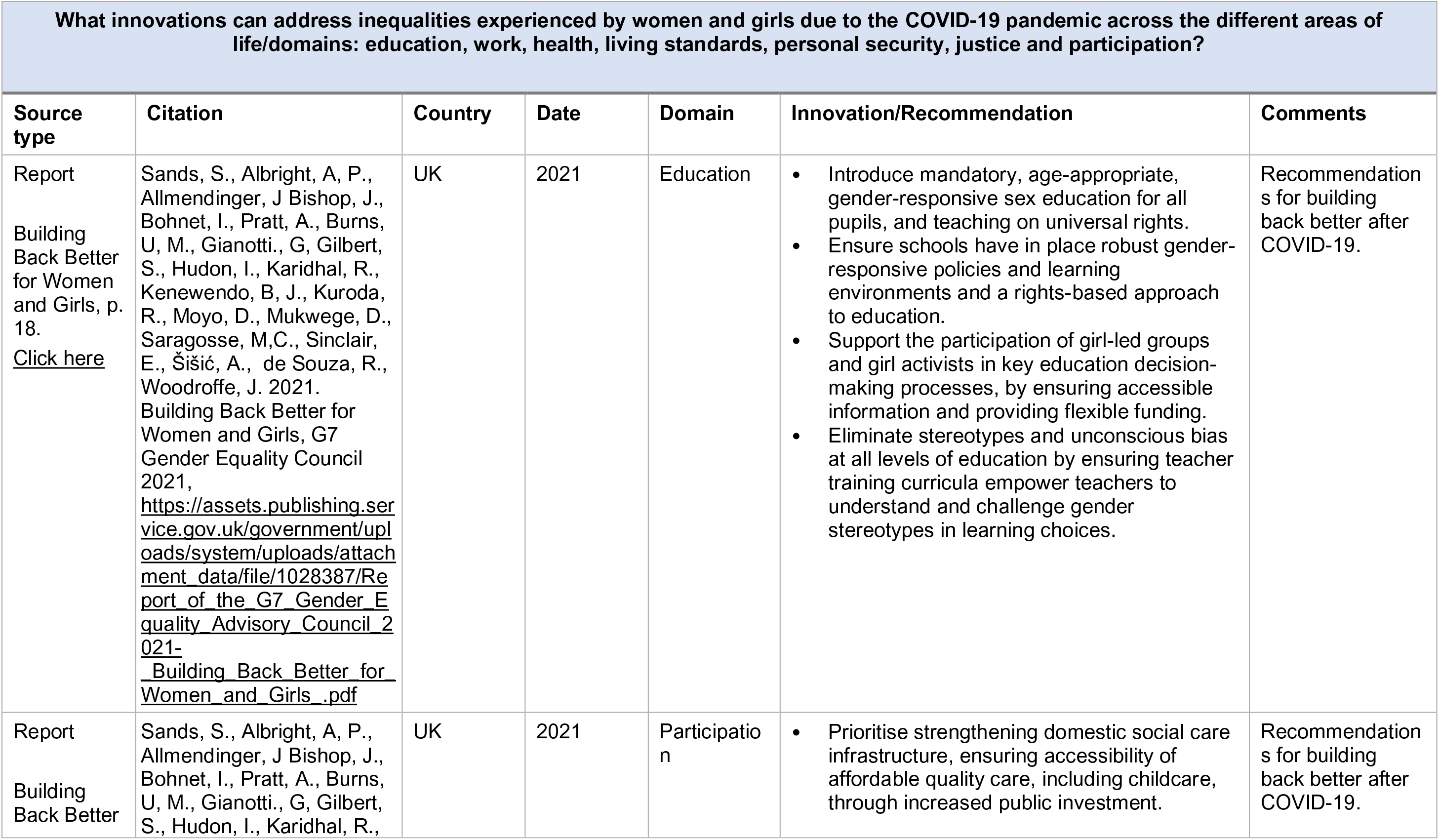
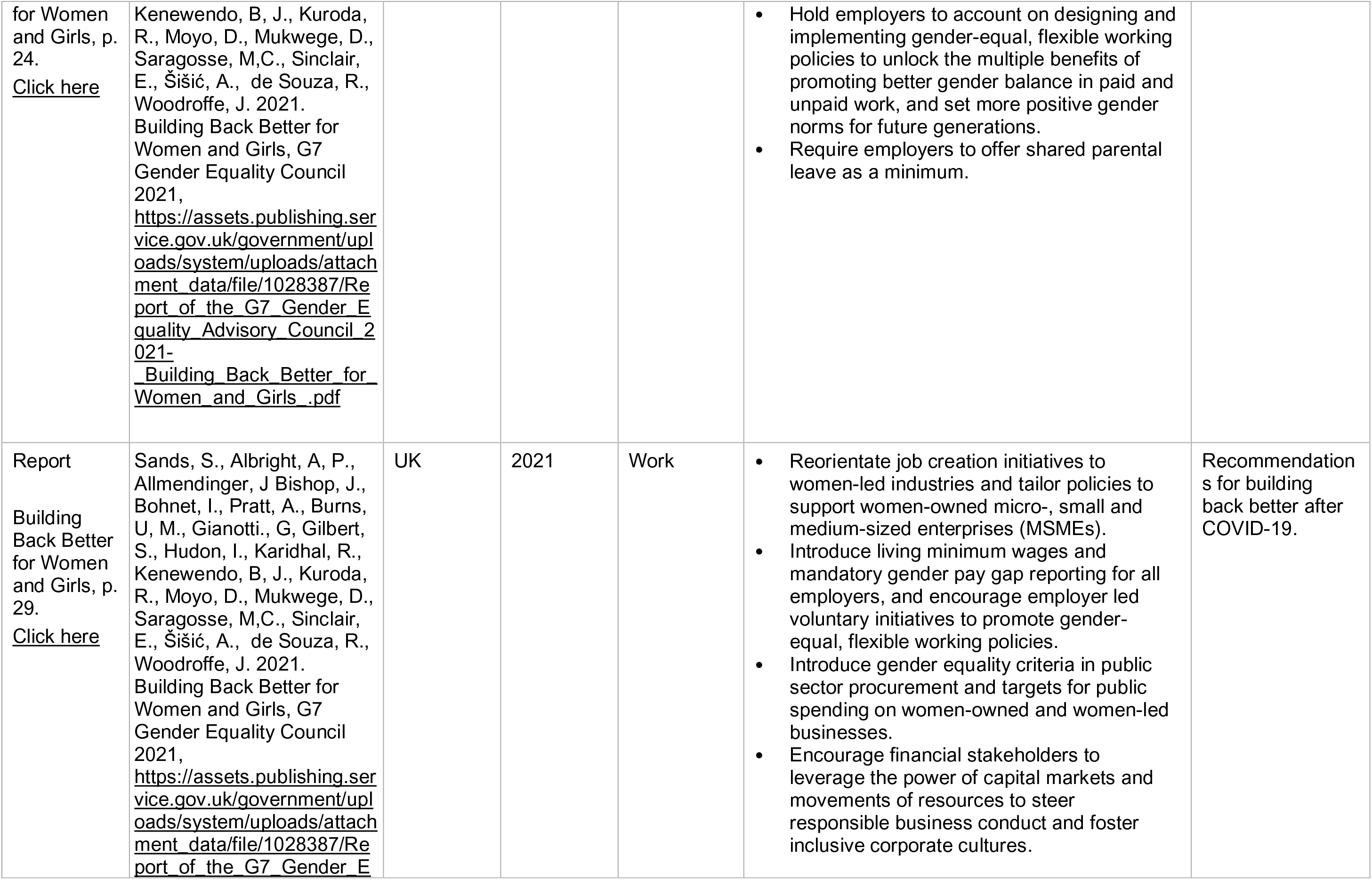
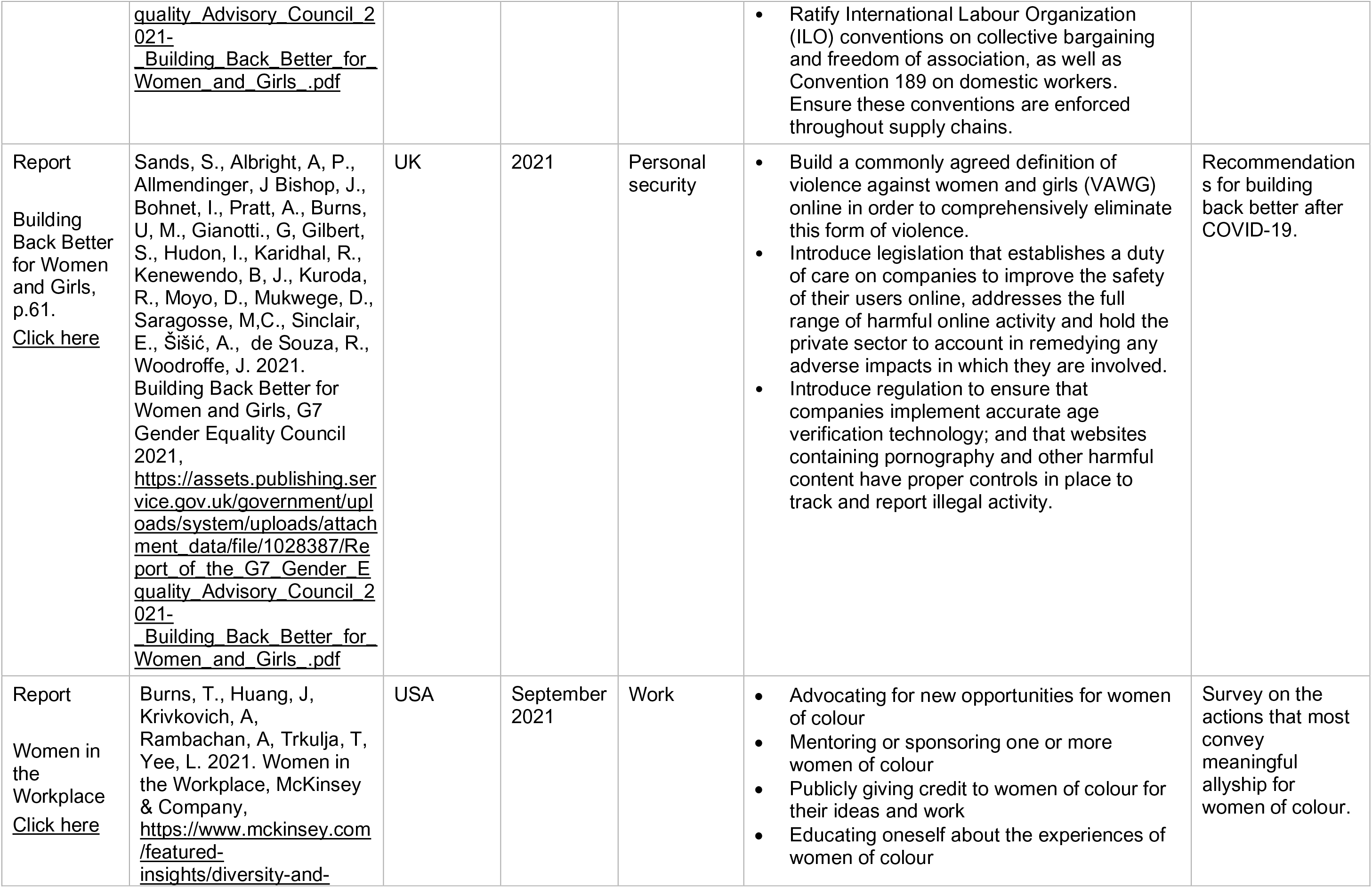
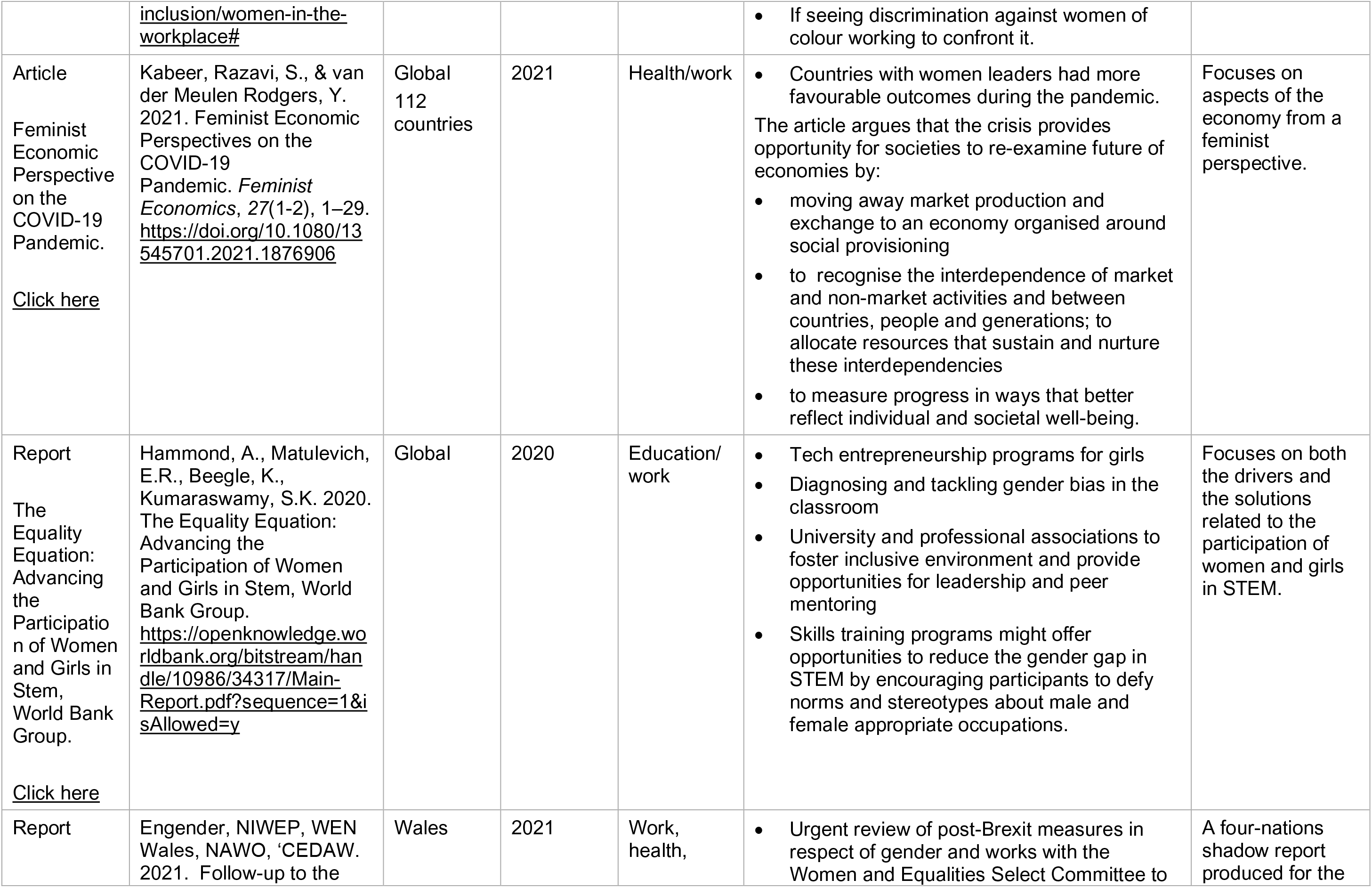
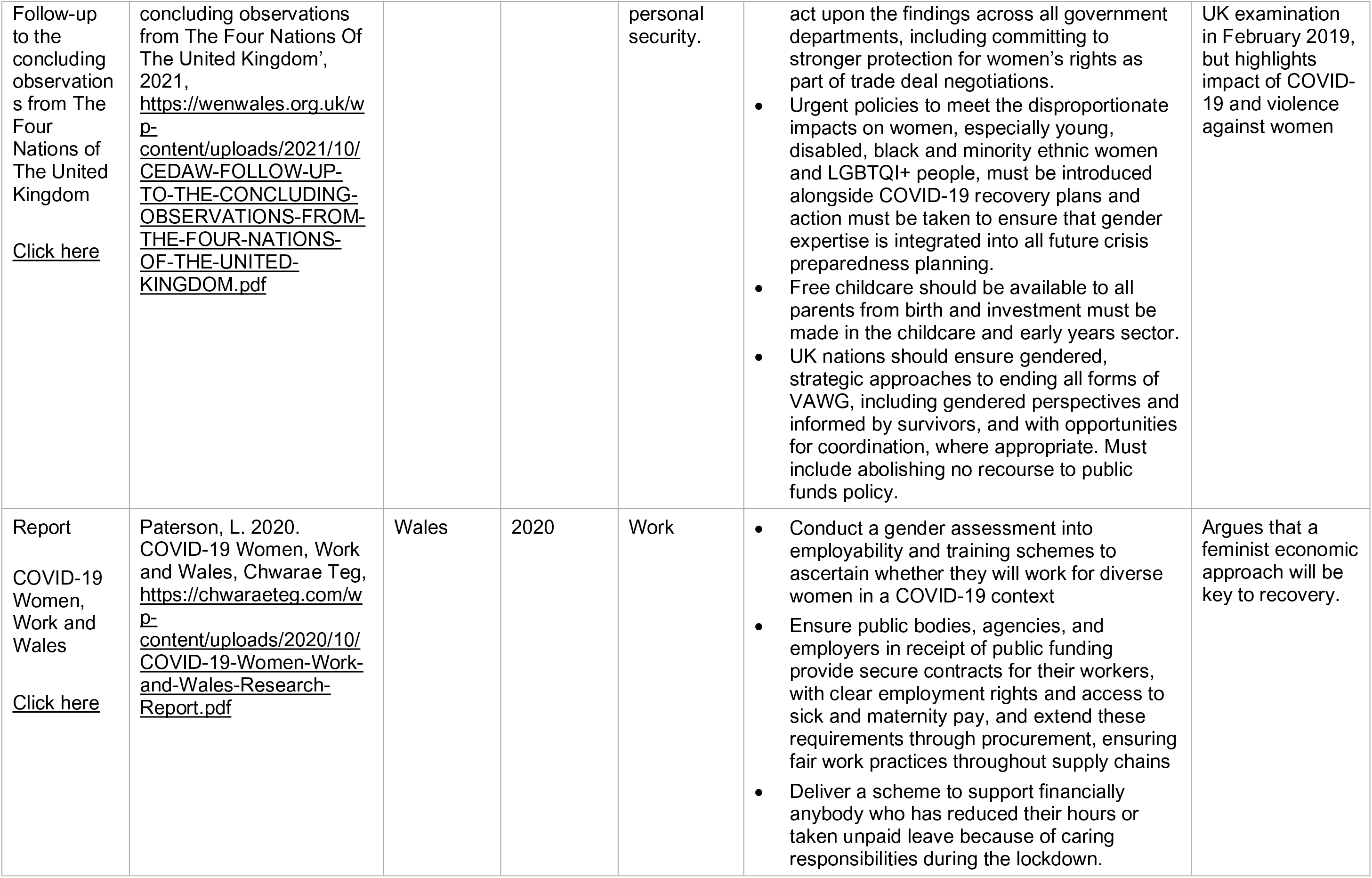
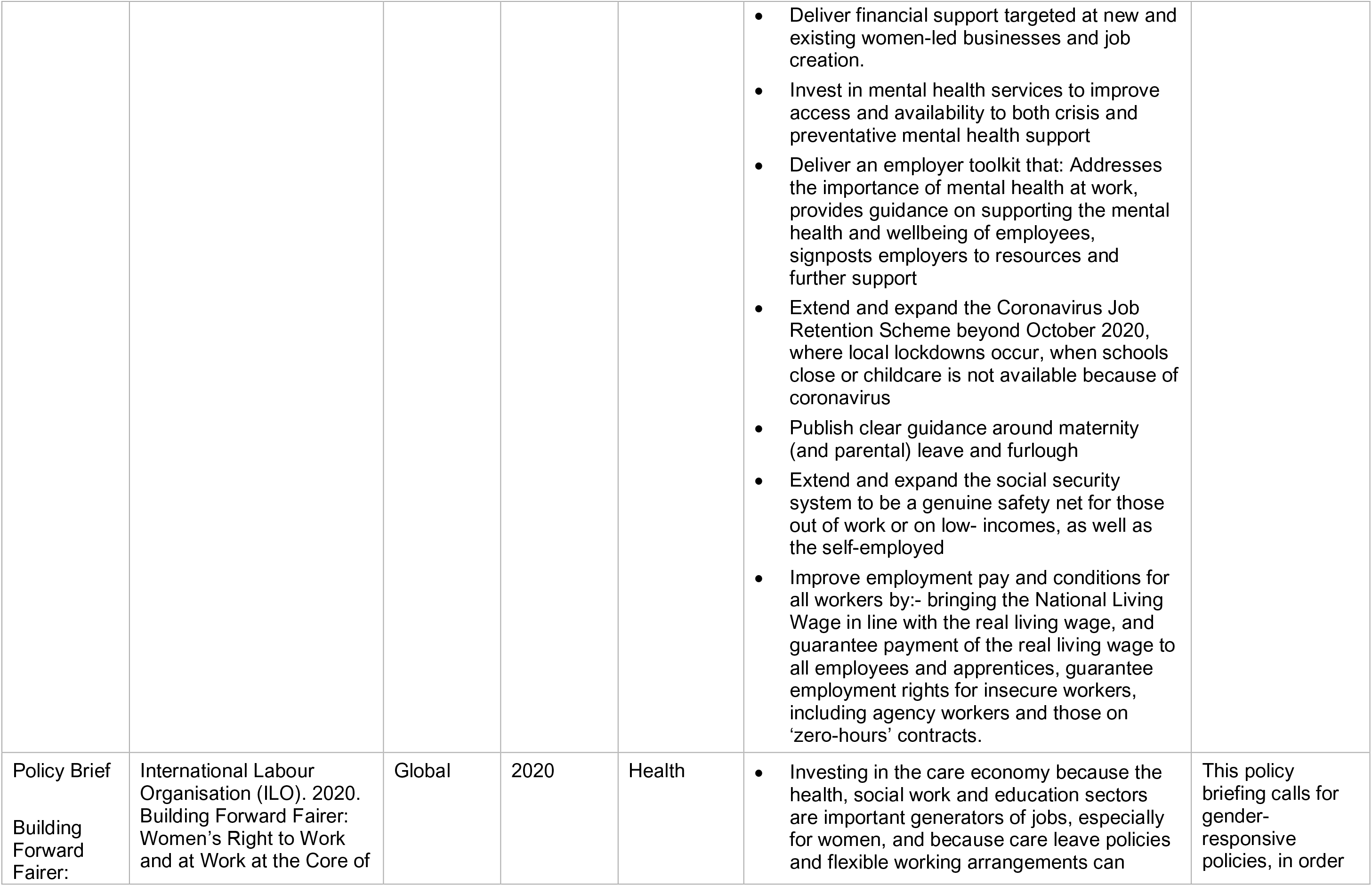
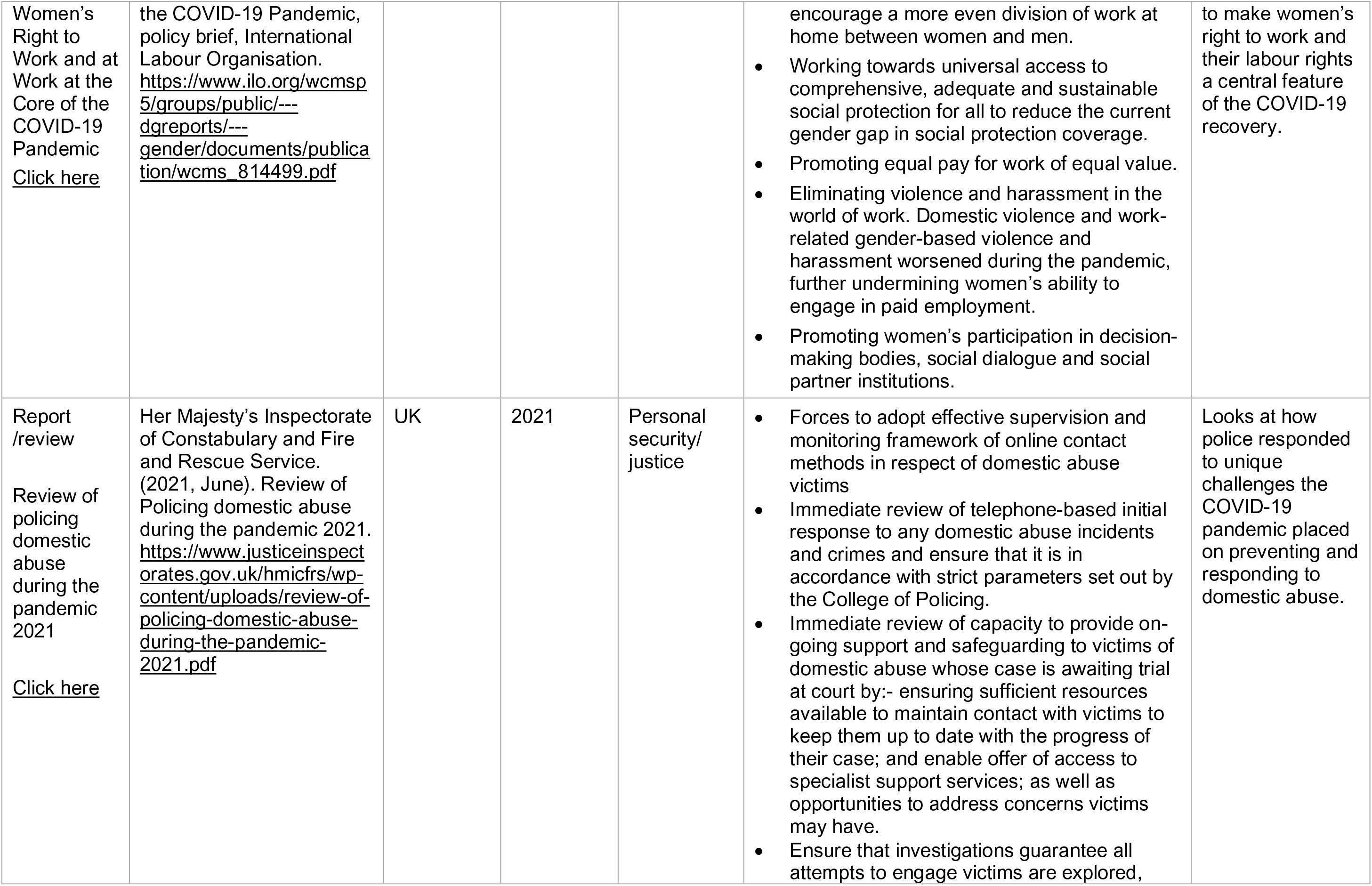
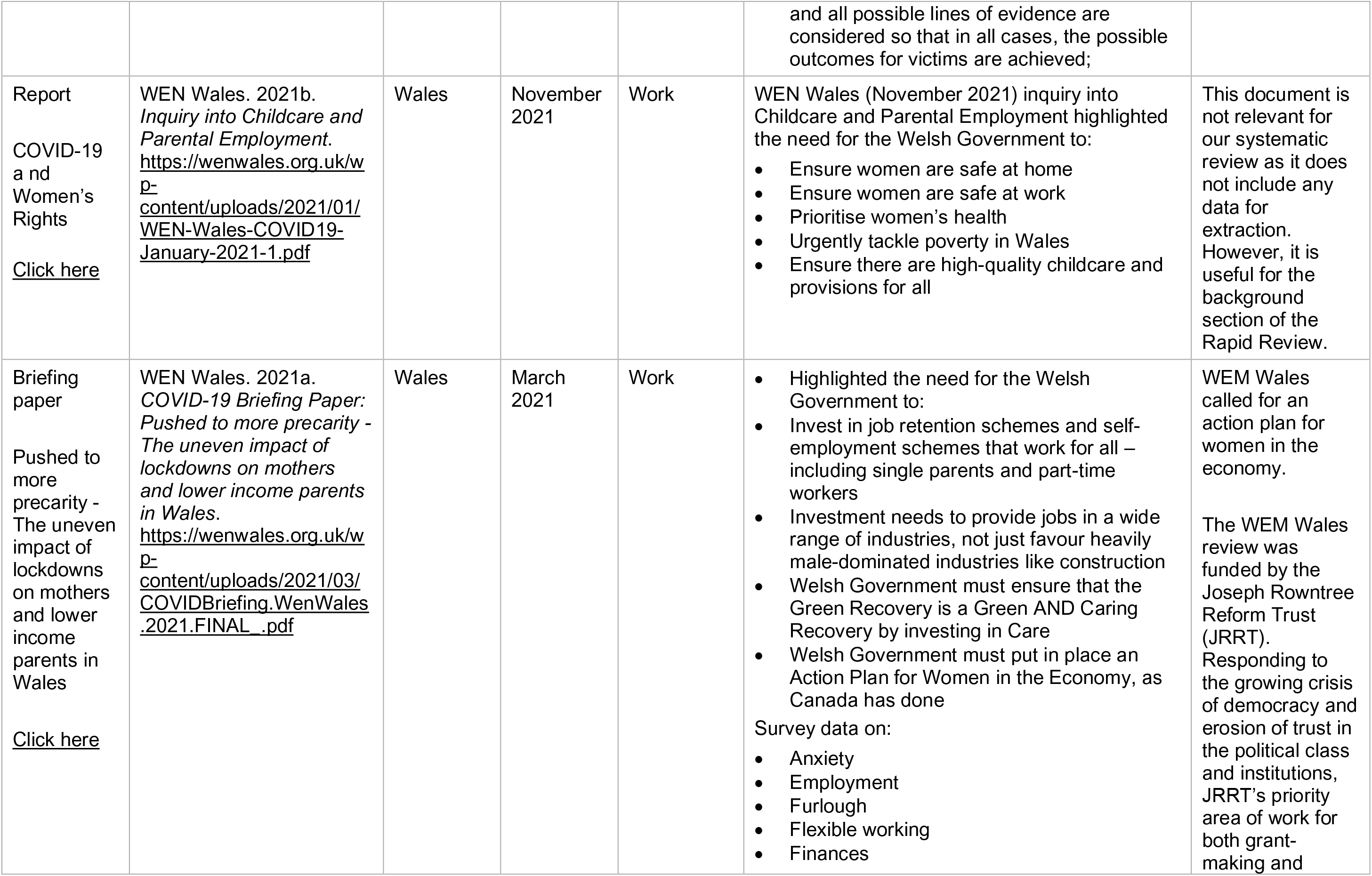
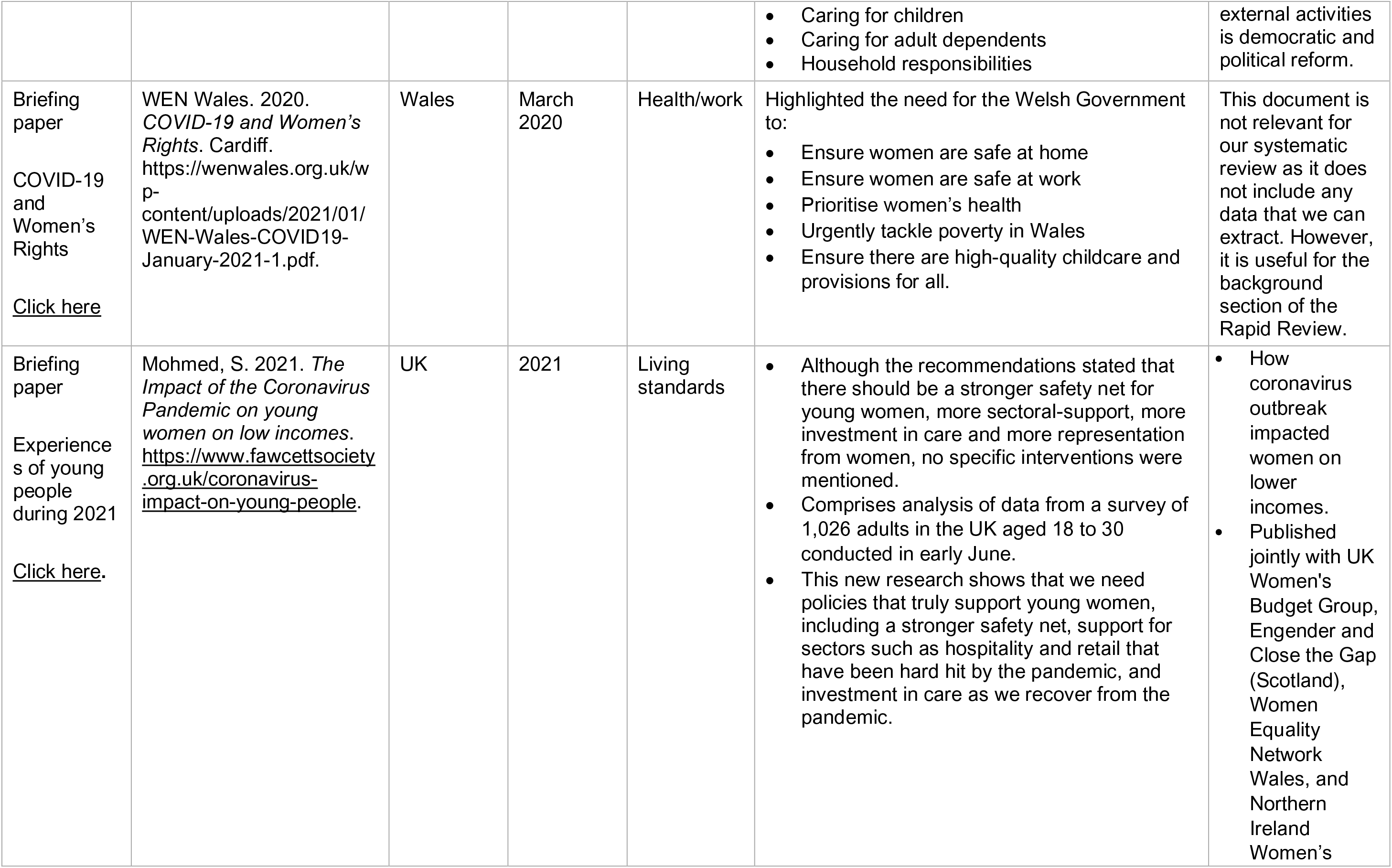
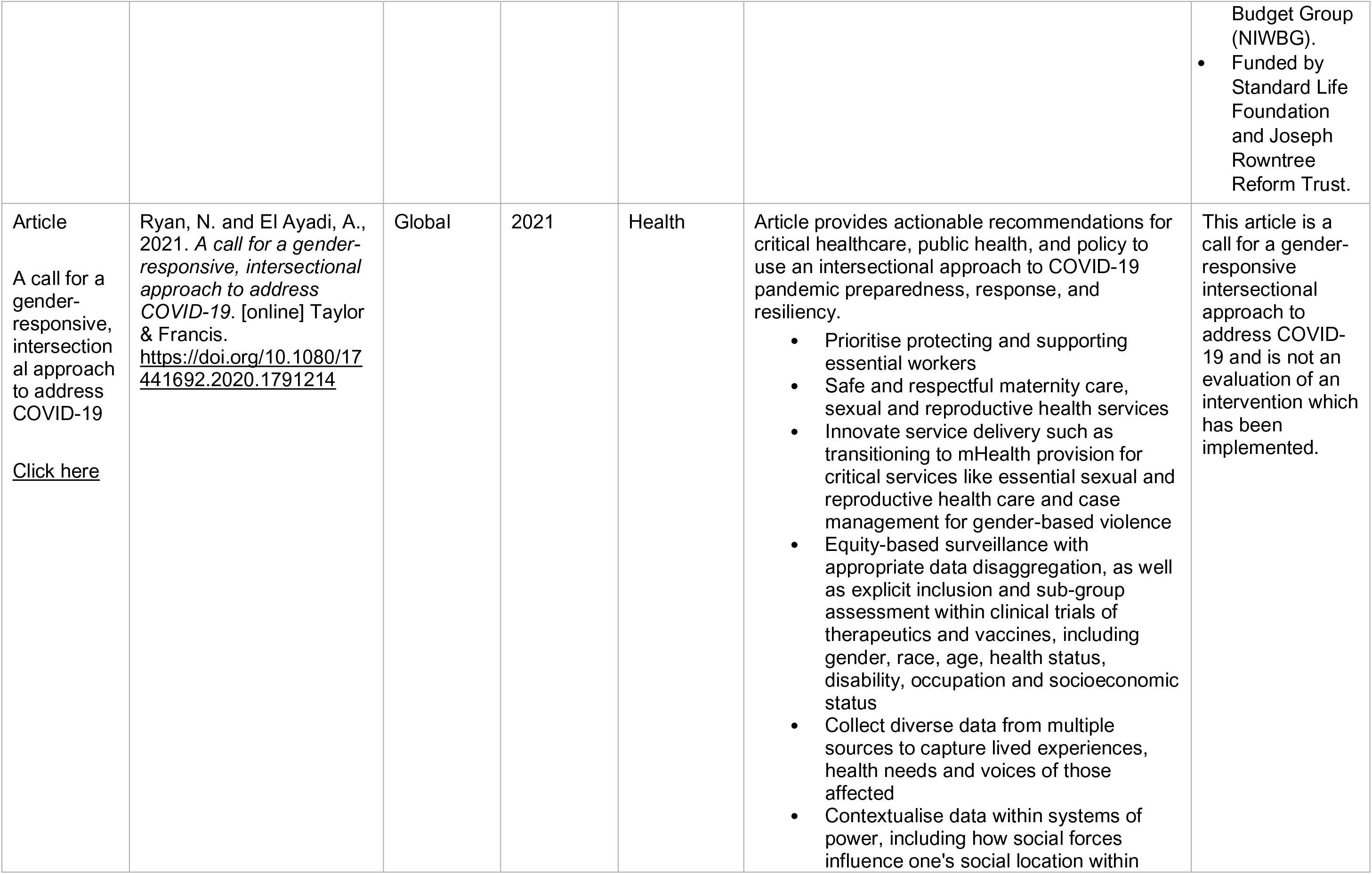
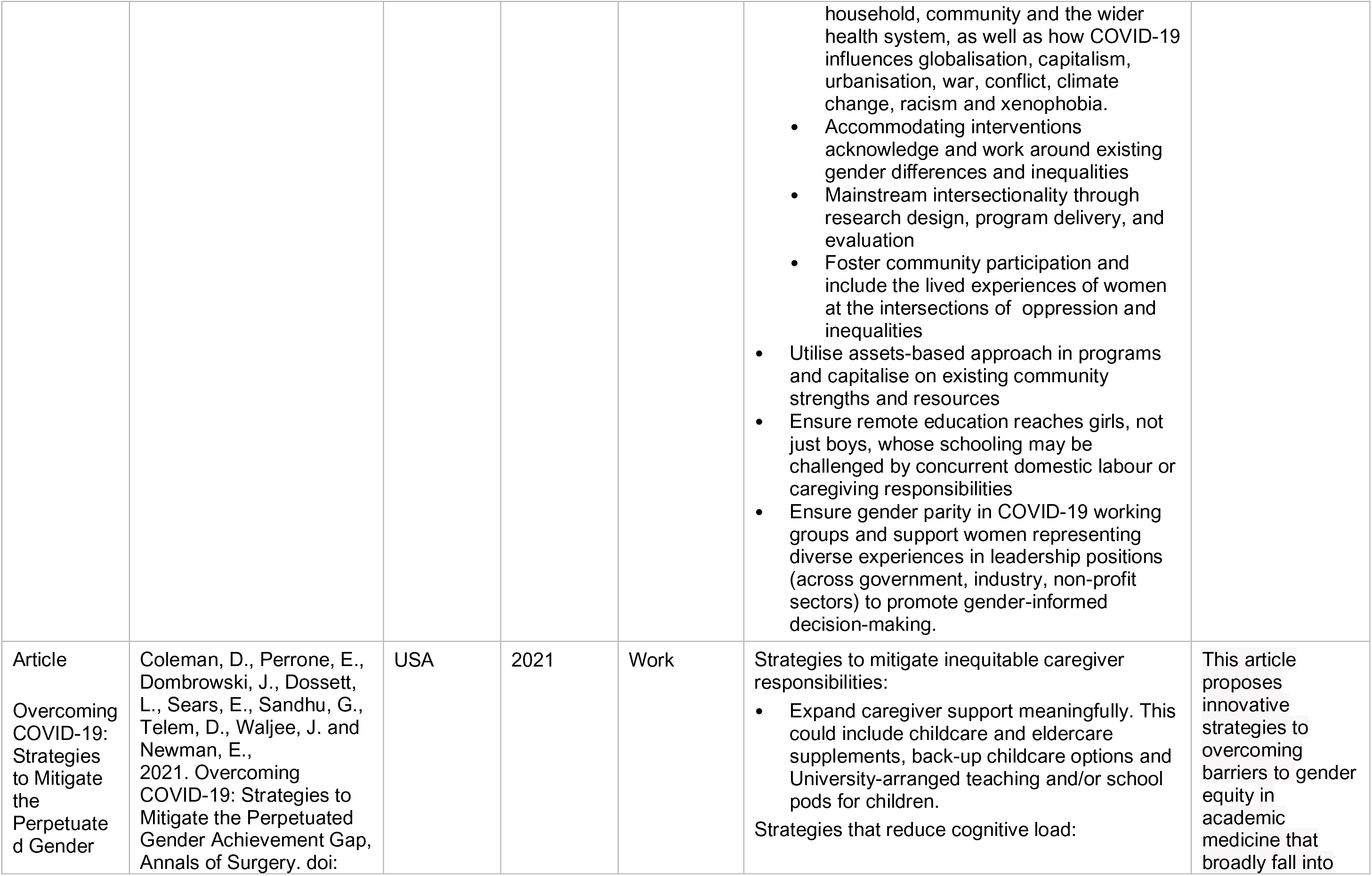
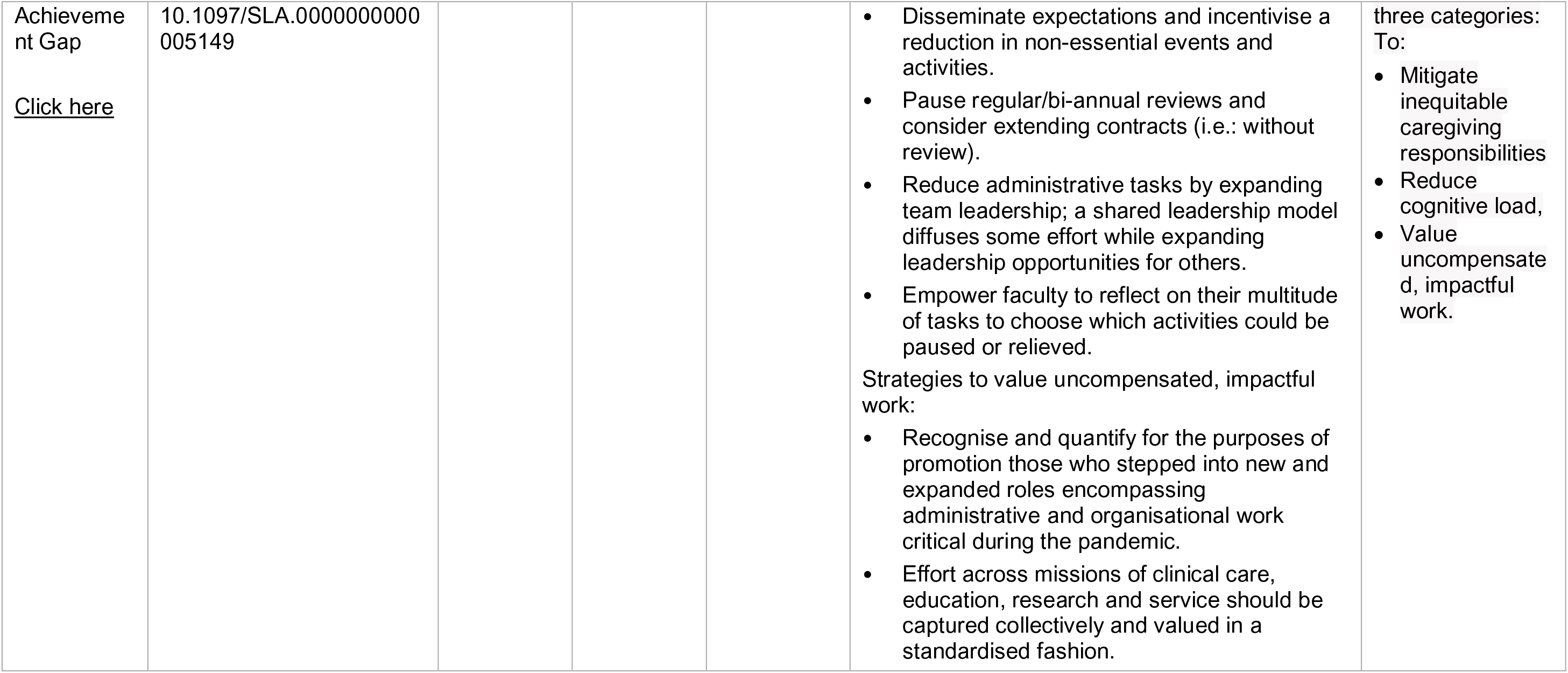

## Abbreviations

AofRCD: Analysis of routinely collected data
Brexit: Brexit is the name given to the United Kingdom’s departure from the European Union. It is a combination of Britain and exit European Union. It is a combination of Britain and exit
BTM: Bridges to Moms
CGD: Center for Global Development
EHRC: Equality and human rights commission
GP: General Practitioner
HCEC: Health and Care Economics Cymru
IPAC: Infection Prevention and Control
JBI: Joanna Briggs Institute
LFD: Lateral Flow Devices
LGBT+: Lesbian, Gay, Bisexual, Transexual, Questioning +. People often use LGBTQ+ to mean all of the “LGBTTTQQIAA” communities including Lesbian, Gay, Bisexual, Transgender, Transsexual, 2/Two-Spirit, Queer, Questioning, Intersex, Asexual, Ally, Pansexual, Agender, Gend Pangender.
LTCF: Long Term Care Facility
M: Mean
NHS: National Health Service
NICE: National Institute for Health and Care Excellence
NS: Unclear or unspecified
OECD: Organisation for Economic Co-operation and Development
PCR: test Polymerase Chain Reaction test
PHE: Public Health England
PHW: Public Health Wales
PICO: framework Participant, Intervention, Comparison, Outcomes framework
PPE: Personal Protective Equipment
RCT: Randomised Controlled Trial
RES: Rapid Evidence Summary
SCIE: Social Care Institute for Excellence
SD: Standard Deviation
SGM: Sexual and Gender Minorities
SR: Suicide Rates
STEM: Science, Technology, Engineering, Mathematics
UK: United Kingdom
VCS: Voluntary and Community Sector
W1: First wave
W2: Second wave
W3: Third wave
WC19EC: Wales COVID-19 Evidence Centre
WHCW: Women Health-Care Workers

